# Single-cell-level spatial transcriptomics identifies erythropoietin-expressing fibroblasts and their tissue niche in human kidney disease

**DOI:** 10.64898/2026.11.26.26369027

**Authors:** Shaun G. Hong, Ara Ko, Jae-Ik Oh, Jung Hun Koh, Donggun Kim, Hyunah Ku, Myeongseop Kim, Youmin Shin, Jeong Ho Joo, Hyun Je Kim, Young-Gon Kim, Jeong Seok Lee, Jinsun Lee, Min Woo Kang, Hajeong Lee, Seung Seok Han, Yaerim Kim, Jinkyung Kwon, Jeonghwan Lee, Jung Pyo Lee, Ji In Park, Jung Tak Park, Jung Nam An, Kyung-Chul Moon, Sehoon Park, Dong Ki Kim, the KORNERSTONE investigators

## Abstract

**Introduction:** Anemia of chronic kidney disease results mainly from insufficient kidney production of erythropoietin (EPO). EPO-producing cells are well characterized in rodents, but little is known about them in the human kidney.

**Methods:** Biopsies from 130 patients with glomerular disease or diabetic kidney disease and 24 living kidney donors were profiled by single-cell-level spatial transcriptomics with 5,101 probes. EPO-expressing fibroblasts were characterized in these sections and in single-nucleus data from our cohort (125 specimens) and the Kidney Precision Medicine Project (157 participants) and in a published single-cell dataset, and their location was examined in two external spatial datasets (209 and 16 participants). Their abundance was related to eGFR, hemoglobin and plasma EPO, measured in 100 patients.

**Results:** Cells with two or more *EPO* transcripts were rare and were predominantly interstitial fibroblasts in tissue sections and single nuclei. In single nuclei, 70 genes were detected more often in EPO-positive than in other fibroblasts, including coagulation factor VIII, *VEGFA*, *GREM2* and prostacyclin synthase; among signaling pathways, hypoxia showed the largest increase in activity and TGF-β the largest decrease. Twenty-one of 29 panel genes and both pathways were reproduced in tissue sections. EPO-expressing cells lay beside intact thick ascending limb, in the zone of straight tubules and away from immune cells. EPO-expressing cells were fewer with proximal tubule injury, which accounted for the association with eGFR, and plasma EPO was higher in patients with more EPO-expressing cells.

**Conclusions:** In human kidney disease, EPO is expressed in the zone of straight tubules by a subset of interstitial fibroblasts with active hypoxia signaling and low TGF-β signaling, a profile reproduced across single-nucleus and spatial datasets. These cells are scarce in injured and inflamed tissue, and their genes provide candidate markers of human EPO-producing cells.

## Introduction

Anemia is a common complication of chronic kidney disease, and its prevalence increases as the glomerular filtration rate declines.^1^ Its main cause is a relative deficiency of erythropoietin (EPO): the plasma EPO concentration does not rise in proportion to the severity of anemia,^2^ and this relative deficiency is present already in the early stages of chronic kidney disease.^3^ Erythropoiesis-stimulating agents replace the hormone, whereas inhibitors of hypoxia-inducible factor (HIF) prolyl hydroxylase stimulate endogenous EPO production^4^ and therefore depend, at least in part, on kidney cells that retain the capacity to produce EPO.^5^

In rodents, EPO is produced by peritubular interstitial fibroblasts,^6^ located mainly in the kidney cortex,^7^ that express HIF-2α.^8^ After kidney injury, these cells cease to produce EPO^9^ as they transdifferentiate into matrix-producing myofibroblasts,^10^ although they can resume EPO production.^11^ Inflammatory cytokines also suppress EPO production in experimental systems.^12^

In contrast, little is known about EPO-producing cells in the human kidney. *EPO* transcripts have been localized to interstitial cells by in situ hybridization in a hypoxic kidney^13^ and in a small series of nephrectomy and biopsy specimens.^14^ The genes and signaling pathways that characterize these cells, their location along the corticomedullary axis and their relation to the surrounding tissue have not been described in patients with kidney disease.

We hypothesized that EPO-expressing cells in the human kidney are interstitial fibroblasts with a distinct molecular profile and tissue location, and that such cells are scarce in injured tissue. We aimed to identify the cells that express EPO in kidney biopsies from patients with glomerular disease or diabetic kidney disease, to characterize their genes, signaling pathways and tissue location, and to relate their abundance to kidney function, hemoglobin and plasma EPO.

## Methods

### Study datasets and participants

The primary dataset consisted of single-cell-level spatial transcriptomics of archival formalin-fixed, paraffin-embedded kidney biopsies from participants of KORNERSTONE, a prospective multicenter Korean kidney biopsy cohort with linked clinical data and biospecimens.^15^ The biopsies were from 130 patients with IgA nephropathy, diabetic kidney disease (DKD), minimal change disease, membranous nephropathy or focal segmental glomerulosclerosis and from 24 living kidney donors who underwent a zero-time biopsy (Table 1). Plasma collected within 30 days of the biopsy was available for 100 of the 130 patients, and plasma EPO was measured in these samples by proximity extension assay (Olink Explore HT)^16^ as described previously.^17^ Independent single-nucleus RNA-sequencing data from the KORNERSTONE cohort comprised 839,765 nuclei, including 25,297 fibroblast nuclei, from 125 kidney specimens obtained at two centers (110 biopsies of the same five diseases and 15 nephrectomy controls) and were processed as described previously.^18^ eGFR at biopsy was calculated with the 2021 CKD-EPI creatinine equation,^19^ hemoglobin was measured at the time of the biopsy, and anemia was defined as a hemoglobin concentration below 13.0 g/dL in males and 12.0 g/dL in females.^20^ No patient was receiving an erythropoiesis-stimulating agent or a HIF prolyl hydroxylase inhibitor at the time of biopsy. The study was conducted in accordance with the Declaration of Helsinki and was approved by the Institutional Review Board of Seoul National University Hospital (No. 2511-060-1690); tissue and plasma were used with written consent for research obtained at biopsy.

**Table 1.** Characteristics of the participants with a spatial section.

| Characteristic | Living donors | IgA nephropathy | DKD | MCD | MN | FSGS |
|---|---|---|---|---|---|---|
| Participants | 24 | 60 | 28 | 24 | 13 | 5 |
| Age, years | 55 (50, 58) | 39 (33, 49) | 51 (38, 59) | 47 (32, 57) | 60 (53, 65) | 28 (21, 43) |
| Female | 17 (71%) | 30 (50%) | 8 (29%) | 9 (38%) | 4 (31%) | 4 (80%) |
| Hemoglobin, g/dL | 13.5 (12.2, 13.9) | 13.6 (12.2, 14.5) | 11.3 (10.0, 12.4) | 14.3 (13.1, 15.6) | 12.8 (12.1, 13.3) | 13.1 (13.1, 13.3) |
| Anemia | 5 (21%) | 11 (18%) | 22 (79%) | 2 (8%) | 5 (38%) | 0 (0%) |
| eGFR, mL/min/1.73 m² | 103 (99, 106) | 93 (79, 112) | 54 (40, 84) | 105 (81, 116) | 103 (97, 106) | 101 (49, 109) |
| UPCR, g/g | 0.05 (0.03, 0.06) | 0.84 (0.57, 1.24) | 4.18 (3.00, 8.22) | 8.33 (4.95, 11.39) | 4.78 (4.07, 7.07) | 6.83 (1.87, 11.91) |
| Tissue area, mm² | 5.6 (4.0, 8.0) | 5.2 (4.1, 6.6) | 3.7 (3.0, 5.4) | 4.6 (2.8, 6.8) | 3.9 (3.0, 5.6) | 6.8 (5.6, 7.8) |
| Fibroblasts | 2,782 (1,897, 3,407) | 5,928 (3,959, 7,942) | 4,641 (3,772, 7,166) | 4,398 (2,012, 6,539) | 3,380 (2,349, 5,548) | 7,945 (5,872, 10,920) |
| Thick ascending limb cells, % of cells | 13.6 (9.5, 21.7) | 13.5 (8.5, 18.9) | 9.4 (7.7, 14.4) | 11.9 (6.3, 18.8) | 11.6 (8.4, 19.3) | 11.0 (8.4, 14.4) |
| Participants with ≥1 EPO-expressing cell, *n* (%) | 12 (50%) | 44 (73%) | 10 (36%) | 8 (33%) | 4 (31%) | 3 (60%) |
| EPO-expressing cells per 1,000 fibroblasts | 0.10 (0.00, 0.85) | 0.30 (0.00, 0.91) | 0.00 (0.00, 0.49) | 0.00 (0.00, 0.43) | 0.00 (0.00, 0.15) | 0.14 (0.00, 0.18) |
| Plasma EPO within 30 days | 0 | 52 | 11 | 23 | 9 | 5 |

Values are median (first quartile, third quartile) or number (percentage). Living donors, zero-time biopsies at transplantation; their age, eGFR and UPCR are the pre-donation values of the donor records. Anemia, hemoglobin below 13.0 g/dL in males and 12.0 g/dL in females. eGFR, 2021 CKD-EPI creatinine equation. EPO-expressing cells, fibroblasts with two or more *EPO* transcripts. Plasma EPO within 30 days, participants with a plasma sample within 30 days of the biopsy; all of them also have a spatial section. Hemoglobin, measured at the time of the biopsy (living donors, at donation). DKD, diabetic kidney disease; eGFR, estimated glomerular filtration rate; EPO, erythropoietin; FSGS, focal segmental glomerulosclerosis; MCD, minimal change disease; MN, membranous nephropathy; UPCR, urine protein-to-creatinine ratio.

### Spatial transcriptomics and cell annotation

Sections were profiled with the Xenium Prime 5K panel (5,101 probes)^21^; tissue processing, cell segmentation and quality control are described in Supplementary Methods. Cells were annotated according to the cell-type hierarchy of the Kidney Precision Medicine Project (KPMP) kidney atlas^22^: the spatial data were integrated with the single-nucleus data from the KORNERSTONE cohort and clustered, and clusters were assigned by canonical markers, with fibroblasts identified by *DCN*, *C7* and *PDGFRA*, and vascular smooth muscle, mesangial and renin-associated cells annotated as separate types (Supplementary Methods). Fibroblasts with two or more *EPO* transcripts were defined as EPO-expressing cells. Each EPO-expressing cell was compared with ten fibroblasts from the same participant that had no *EPO* transcript and a similar library size, and the comparison was repeated with fibroblasts from the same tissue zone (Supplementary Methods).

### External validation datasets

Four published datasets were used: the KPMP single-nucleus RNA sequencing dataset (615,169 nuclei from 157 participants enrolled with chronic kidney disease, acute kidney injury or resilient diabetes, or as healthy references),^22^ a single-cell RNA sequencing dataset of PDGFRβ-positive kidney cells (30,270 cells from 8 patients),^23^ the KPMP spatial transcriptomics dataset (Visium; 210 samples from 209 participants) and an external Xenium dataset (16 participants: 8 with DKD, 5 with other kidney diseases and 3 controls).^24^ Participant characteristics are given in Supplementary Table S1.

### Genes and signaling pathways

The genes associated with EPO expression were examined across the whole transcriptome in the single-nucleus data from the KORNERSTONE cohort and in two external resources, the KPMP single-nucleus dataset and the dataset of PDGFRβ-positive cells. Counts are sparser in these datasets than in tissue sections, and nuclei or cells with one or more *EPO* counts were therefore defined as EPO-positive. The frequency of EPO-positive nuclei was compared between fibroblasts and other nuclei by a rate ratio, and EPO-positive fibroblast nuclei were compared with matched fibroblast nuclei from the same participant; estimates were combined across the datasets. The genes detected more often in EPO-positive nuclei were tested for over-representation of Gene Ontology biological processes,^25^ Reactome pathways^26^ and imprinted genes. In both the single-nucleus and the tissue comparison, the activity of the 14 signaling pathways of the PROGENy model was inferred from the genes that respond to each pathway,^27^ and Hallmark gene sets^28^ were compared by rank; in the nuclei, transcription factor activity was inferred from target genes. A profile score, defined as the mean expression of the genes detected more often in EPO-positive nuclei (for KPMP, the genes identified in the other two datasets), was calculated for every fibroblast nucleus of KPMP and every fibroblast of the spatial dataset (Supplementary Methods).

### Neighborhoods, tissue squares and zones

The neighborhood of a fibroblast was defined as its 20 nearest cells within 30 µm. Within biopsies, the number of EPO-expressing cells per 50-µm tissue square was related to the proportion of immune cells, the contractile program of the surrounding fibroblasts (an expression program led by *ACTA2*), the inflammatory signaling of these fibroblasts (mean activity of the TNF-α, NF-κB and JAK–STAT pathways) and the proportion of proximal tubule cells that were injured, by Poisson models with participant fixed effects. Squares were assigned to the cortical labyrinth, the zone of straight tubules or the deeper medulla according to the tubular segments in the surrounding 3 × 3 block of squares (150 µm), and the assignment was repeated without the thick ascending limb. Clustering of EPO-expressing cells was compared with 1,000 random reassignments of EPO expression among the fibroblasts of the same biopsy and zone.

### Statistical analysis

The number of EPO-expressing cells per participant was modeled by negative binomial regression with the number of fibroblasts as offset, adjusted for age, sex, eGFR and the proportion of thick ascending limb cells; diagnosis was added when diagnoses were combined. The association of plasma EPO with the density of EPO-expressing cells was estimated by least-squares regression with heteroskedasticity-consistent standard errors, adjusted for age, sex, eGFR, assay plate and diagnosis. Estimates are reported with 95% confidence intervals. Multiple testing within each family of tests was controlled by the Benjamini–Hochberg procedure.^29^

## Results

### Datasets

The spatial dataset comprised biopsies from 130 patients and 24 living kidney donors (Table 1). Most patients had preserved kidney function (104 had an eGFR of 60 mL/min/1.73 mZ or higher); patients with DKD had the lowest eGFR and were most often anemic. EPO-positive cells were also examined in three single-cell datasets and two external spatial datasets, whose participants are described in Supplementary Table S1.

### *EPO* transcripts are found mainly in interstitial fibroblasts

In the spatial dataset, two thirds of the cells with two or more *EPO* transcripts were fibroblasts (684 of 1,031; Figure 1a,b), about 1 in 1,000 fibroblasts. Of the 2,251 fibroblasts with an *EPO* transcript, 684 (30%) had two or more. For the 40 negative control probes of the panel, the same proportion was 1% (11 of 1,100, summed over the probes; *P* < 0.001; Figure 1b; Supplementary Table S1; Supplementary Data 1). Nearly half of the 347 other cells with two or more *EPO* transcripts were in contact with an EPO-expressing fibroblast, compared with 0.4% of matched cells of the same types (Supplementary Table S1). A single transcript was found at a similarly low frequency in every cell type and was not counted as expression (Supplementary Results; Supplementary Figure S1a). In the single-nucleus data from the KORNERSTONE cohort and from KPMP, nuclei with an *EPO* count were three to four times as frequent among fibroblasts as among other nuclei (Figure 1c,d; Supplementary Table S1), and most nuclei with two or more counts were fibroblasts (27 of 37). Most cells with two or more counts in the external Xenium dataset were also fibroblasts (172 of 241; Figure 1e). EPO-expressing cells were present in 81 of the 154 biopsies, including half of the biopsies from living donors. Nearly two thirds of the EPO-expressing cells had another EPO-expressing cell within 30 µm, against 4% when the same number of cells was placed at random among the fibroblasts of the same biopsy and zone (*P* < 0.001; Supplementary Table S1).

**Figure 1.**
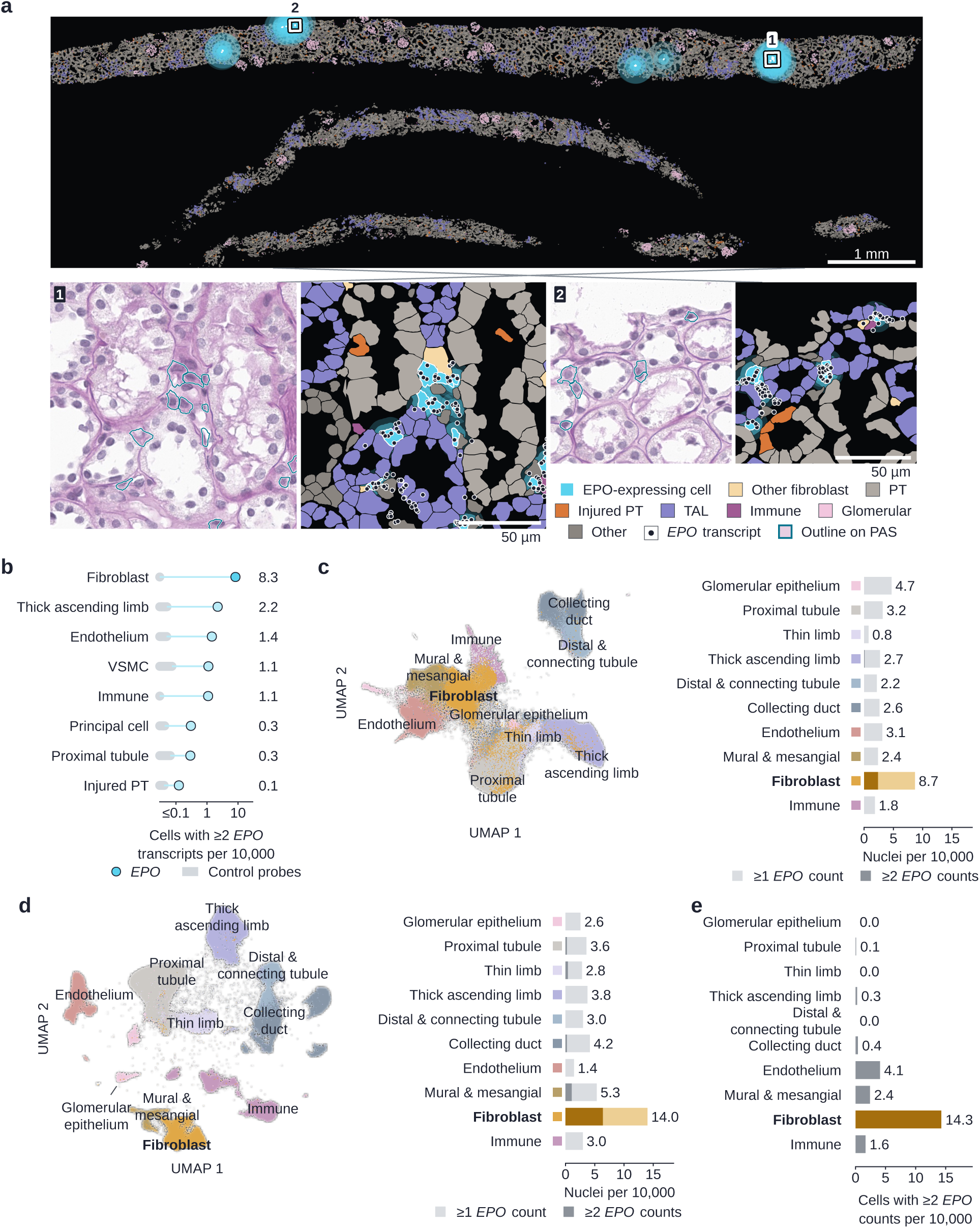
EPO transcripts are most frequent in interstitial fibroblasts in each dataset. **a**, Representative section of the spatial dataset with every segmented cell filled by class and EPO-expressing cells marked; boxes, the windows shown beside the section on periodic acid–Schiff (PAS; outlines, EPO-expressing cells) and as segmented cells (points, *EPO* transcripts); scale bars, 1 mm and 50 µm. **b**, Cells with two or more *EPO* transcripts per 10,000 cells by cell type in the spatial dataset; gray band, the negative control probes of the panel (mean to maximum). **c**, Single-nucleus data from the KORNERSTONE cohort: embedding of all nuclei colored by cell class, with the classes named on the embedding, and nuclei with *EPO* counts per 10,000 nuclei by cell class (light, one or more counts; dark, two or more). **d**, KPMP single-nucleus dataset, as in **c**. **e**, External Xenium dataset: cells with two or more *EPO* counts per 10,000 cells by cell class. KPMP, Kidney Precision Medicine Project; PT, proximal tubule; TAL, thick ascending limb; UMAP, uniform manifold approximation and projection; VSMC, vascular smooth muscle cell.

### EPO-positive fibroblasts share a set of genes across datasets

Across the three single-cell datasets, 70 genes were detected more often in EPO-positive fibroblast nuclei than in other fibroblast nuclei from the same participants (Table 2; Supplementary Figure S2a; Supplementary Data 2). The largest differences were found for coagulation factor VIII (*F8*), *VEGFA*, the secreted bone morphogenetic protein antagonist *GREM2*, prostacyclin synthase (*PTGIS*) and the cyclic AMP phosphodiesterase *PDE8B* (Figure 2a). Most of the 70 genes were not included in published hypoxia gene sets or marker lists of EPO-producing cells. These genes were reproduced in tissue sections: of the 29 genes that could be tested in the spatial dataset, 21 were detected more often in the 684 EPO-expressing cells than in other fibroblasts of the same biopsy and none less often (*F8* in 15% versus 0.3% of cells and *PTGIS* in 32% versus 2%), and transcripts of *F8*, *GREM2*, *PTGIS* and *VEGFA* were located within EPO-expressing cells in the sections (Figure 2b; Supplementary Figure S2b; Supplementary Table S2). When the fibroblast nuclei of KPMP were scored with the genes identified in the other two datasets, EPO-positive nuclei ranked among the highest (median, 94th percentile; Figure 2c).

**Figure 2.**
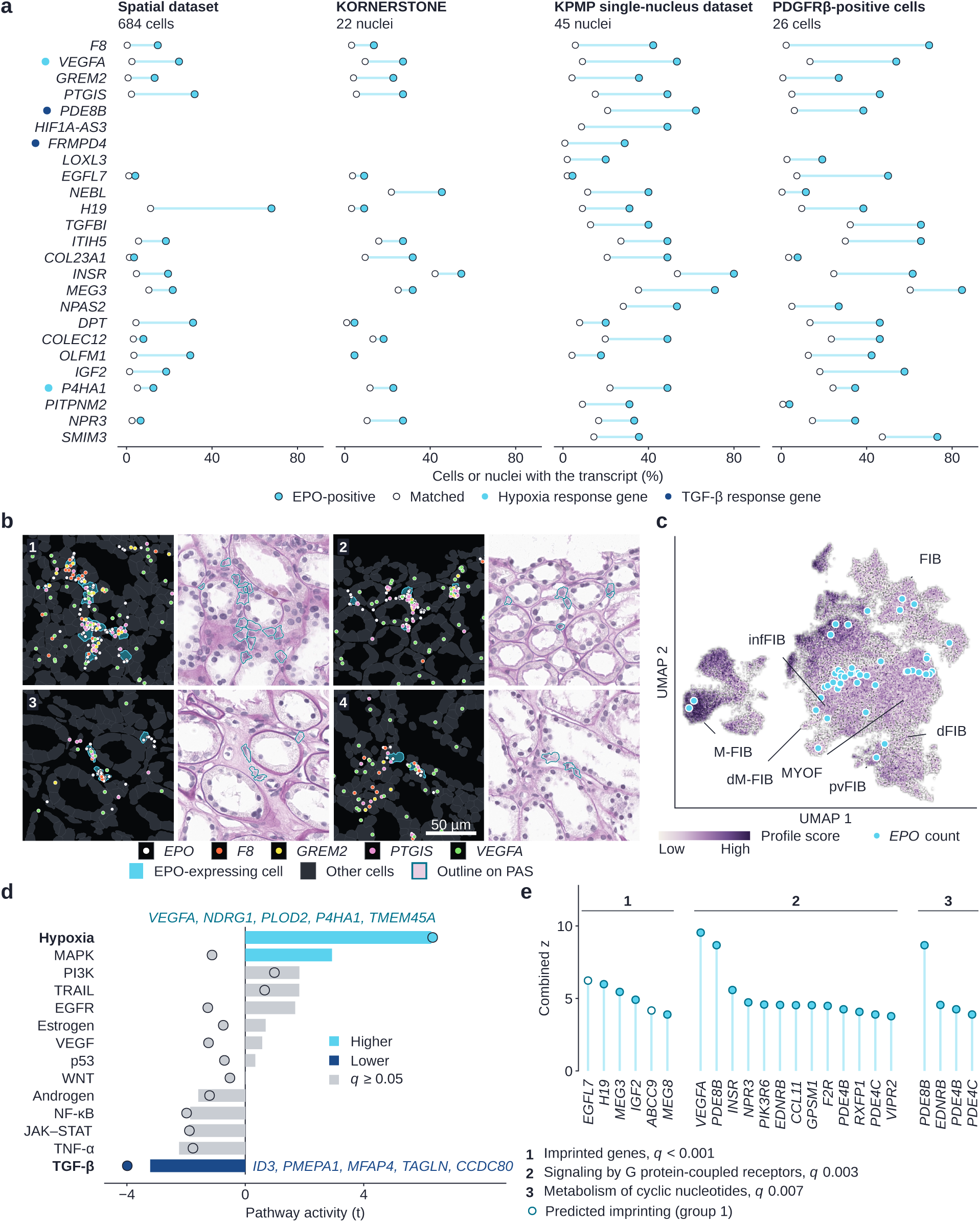
EPO-positive fibroblasts show higher hypoxia signaling and lower TGF-β signaling. **a**, Detection of the 25 genes of Table 2 in EPO-positive cells or nuclei (filled) and in matched cells or nuclei from the same participants (open) in the spatial dataset and in the three single-cell datasets; marks beside the gene names, response genes of the hypoxia and of the TGF-β pathway. **b**, Representative 150-µm windows with EPO-expressing cells, shown as segmented cells (cyan, EPO-expressing cells; points, transcripts of *EPO*, *F8*, *GREM2*, *PTGIS* and *VEGFA*) and on PAS; scale bar, 50 µm. **c**, Embedding of the 32,863 fibroblast nuclei of KPMP colored by the profile score built from the genes of the other two datasets; cyan points, the 46 nuclei with an *EPO* count; names, subclasses, each joined to its region. **d**, Activity of the 14 signaling pathways of the PROGENy model in EPO-positive versus matched fibroblast nuclei (bars) and in EPO-expressing versus matched fibroblasts in tissue sections (points), scored by the genes that respond to each pathway; colored, *q* < 0.05; names, the five response genes that contributed most to the higher hypoxia activity and to the lower TGF-β activity in the nuclei. **e**, Genes of the three enriched groups named in the text, with a gene shown in each of its groups; height, combined *z*; open points in group 1, genes predicted to be imprinted. dFIB, degenerative fibroblast; dM-FIB, degenerative medullary fibroblast; FIB, fibroblast; infFIB, inflammatory fibroblast; KPMP, Kidney Precision Medicine Project; M-FIB, medullary fibroblast; MYOF, myofibroblast; pvFIB, perivascular fibroblast; UMAP, uniform manifold approximation and projection.

**Table 2.**
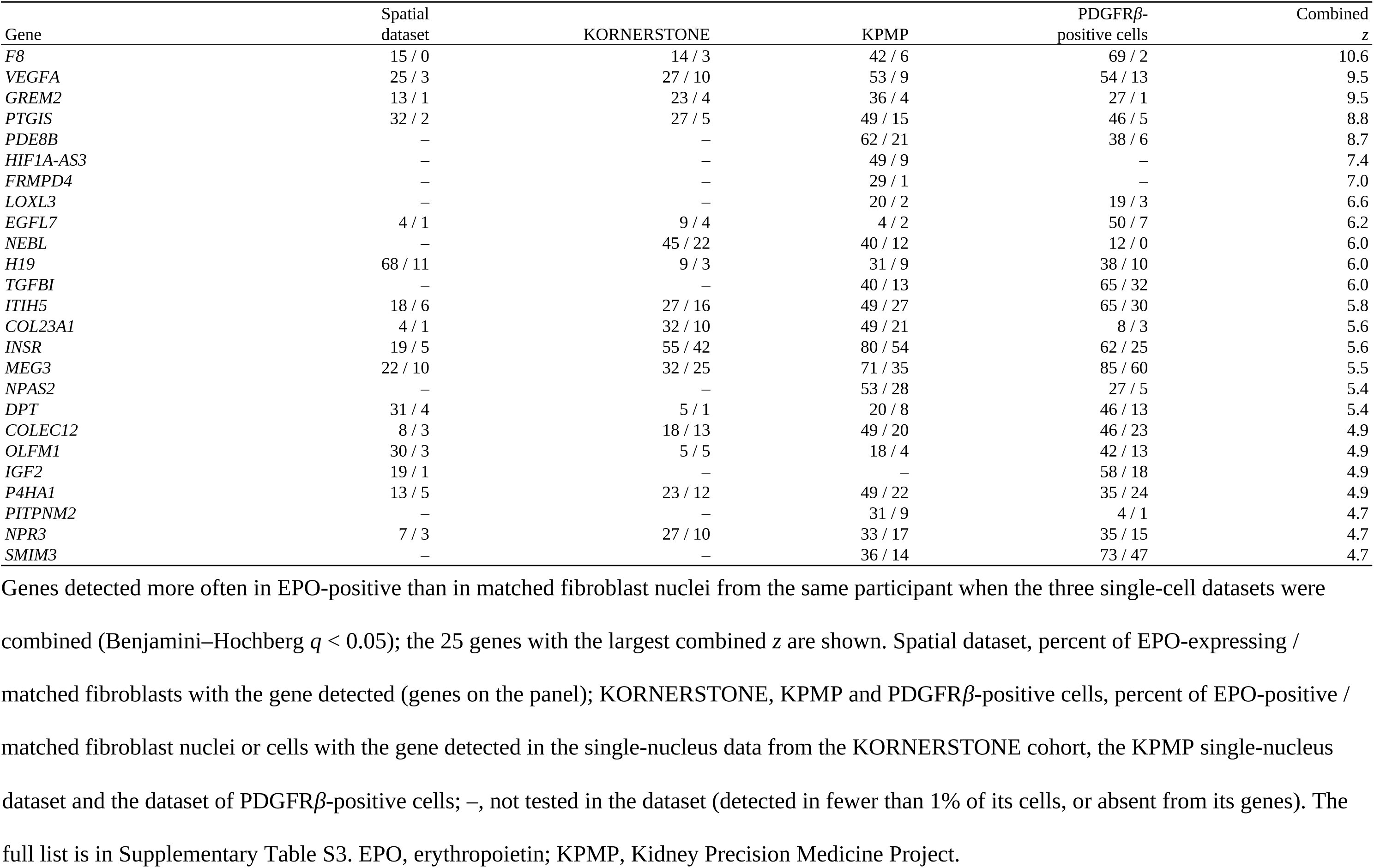
Genes of EPO-positive fibroblasts.

| Gene | Spatial dataset | KORNERSTONE | KPMP | PDGFR*β*-positive cells | Combined *z* |
|---|---|---|---|---|---|
| *F8* | 15 / 0 | 14 / 3 | 42 / 6 | 69 / 2 | 10.6 |
| *VEGFA* | 25 / 3 | 27 / 10 | 53 / 9 | 54 / 13 | 9.5 |
| *GREM2* | 13 / 1 | 23 / 4 | 36 / 4 | 27 / 1 | 9.5 |
| *PTGIS* | 32 / 2 | 27 / 5 | 49 / 15 | 46 / 5 | 8.8 |
| *PDE8B* | – | – | 62 / 21 | 38 / 6 | 8.7 |
| *HIF1A-AS3* | – | – | 49 / 9 | – | 7.4 |
| *FRMPD4* | – | – | 29 / 1 | – | 7.0 |
| *LOXL3* | – | – | 20 / 2 | 19 / 3 | 6.6 |
| *EGFL7* | 4 / 1 | 9 / 4 | 4 / 2 | 50 / 7 | 6.2 |
| *NEBL* | – | 45 / 22 | 40 / 12 | 12 / 0 | 6.0 |
| *H19* | 68 / 11 | 9 / 3 | 31 / 9 | 38 / 10 | 6.0 |
| *TGFBI* | – | – | 40 / 13 | 65 / 32 | 6.0 |
| *ITIH5* | 18 / 6 | 27 / 16 | 49 / 27 | 65 / 30 | 5.8 |
| *COL23A1* | 4 / 1 | 32 / 10 | 49 / 21 | 8 / 3 | 5.6 |
| *INSR* | 19 / 5 | 55 / 42 | 80 / 54 | 62 / 25 | 5.6 |
| *MEG3* | 22 / 10 | 32 / 25 | 71 / 35 | 85 / 60 | 5.5 |
| *NPAS2* | – | – | 53 / 28 | 27 / 5 | 5.4 |
| *DPT* | 31 / 4 | 5 / 1 | 20 / 8 | 46 / 13 | 5.4 |
| *COLEC12* | 8 / 3 | 18 / 13 | 49 / 20 | 46 / 23 | 4.9 |
| *OLFM1* | 30 / 3 | 5 / 5 | 18 / 4 | 42 / 13 | 4.9 |
| *IGF2* | 19 / 1 | – | – | 58 / 18 | 4.9 |
| *P4HA1* | 13 / 5 | 23 / 12 | 49 / 22 | 35 / 24 | 4.9 |
| *PITPNM2* | – | – | 31 / 9 | 4 / 1 | 4.7 |
| *NPR3* | 7 / 3 | 27 / 10 | 33 / 17 | 35 / 15 | 4.7 |
| *SMIM3* | – | – | 36 / 14 | 73 / 47 | 4.7 |

Genes detected more often in EPO-positive than in matched fibroblast nuclei from the same participant when the three single-cell datasets were combined (Benjamini–Hochberg *q* < 0.05); the 25 genes with the largest combined *z* are shown. Spatial dataset, percent of EPO-expressing / matched fibroblasts with the gene detected (genes on the panel); KORNERSTONE, KPMP and PDGFR*β*-positive cells, percent of EPO-positive / matched fibroblast nuclei or cells with the gene detected in the single-nucleus data from the KORNERSTONE cohort, the KPMP single-nucleus dataset and the dataset of PDGFR*β*-positive cells; –, not tested in the dataset (detected in fewer than 1% of its cells, or absent from its genes). The full list is in Supplementary Table S3. EPO, erythropoietin; KPMP, Kidney Precision Medicine Project.

### Hypoxia signaling is higher and TGF-β signaling lower in EPO-positive fibroblasts

*EPAS1*, which encodes HIF-2α, was detected more often in EPO-positive nuclei than in other fibroblast nuclei, and so were the response genes of the hypoxia pathway, the largest difference among the 14 pathways of the PROGENy model (*q* < 0.001); the response genes of the TGF-β pathway were detected less often, the largest difference in the other direction (q 0.009; Figure 2d; Supplementary Table S3). In tissue sections, *EPAS1* was likewise detected more often in EPO-expressing cells than in other fibroblasts of the same biopsy (78% versus 40% of cells). *ACTA2*, *MYLK*, *TNC* and *PDGFRB* were detected less often in EPO-expressing cells, whereas *COL1A1* did not differ, and the Hallmark epithelial–mesenchymal transition set ranked lowest. In tissue sections the same two pathways differed most (*q* < 0.001 each; Figure 2d). These differences persisted when EPO-expressing cells were compared with fibroblasts of the same tissue zone: 20 of the 21 genes shared by nuclei and tissue sections remained more frequently detected, hypoxia-pathway activity remained higher and TGF-β-pathway activity remained lower (Supplementary Tables S2 and S3). In the external Xenium dataset, *EPAS1* and *VEGFA* were also detected more often in fibroblasts with *EPO* transcripts than in other fibroblasts (Supplementary Figure S2c).

The 70 genes were enriched for imprinted genes (*H19*, *IGF2*, *MEG3* and *MEG8*), for G protein-coupled receptor signaling (receptors for endothelin, thrombin, relaxin and vasoactive intestinal peptide) and for cyclic nucleotide metabolism (the cyclic AMP phosphodiesterases *PDE8B*, *PDE4B* and *PDE4C*; Figure 2e; Supplementary Table S3).

### EPO-expressing cells lie beside intact thick ascending limb in the zone of straight tubules

EPO-expressing cells were surrounded by thick ascending limb: thick ascending limb cells were three times as frequent among their neighbors as among the neighbors of other fibroblasts (33% versus 10%), and nearly all adjacent tubular cells were in a healthy state (Figure 3a,b; Supplementary Figure S3a–c; Supplementary Tables S4 and S5; Supplementary Data 3). Proximity to the thick ascending limb was also observed in the KPMP spatial transcriptomics dataset and in the external Xenium dataset (Supplementary Figure S3d).

**Figure 3.**
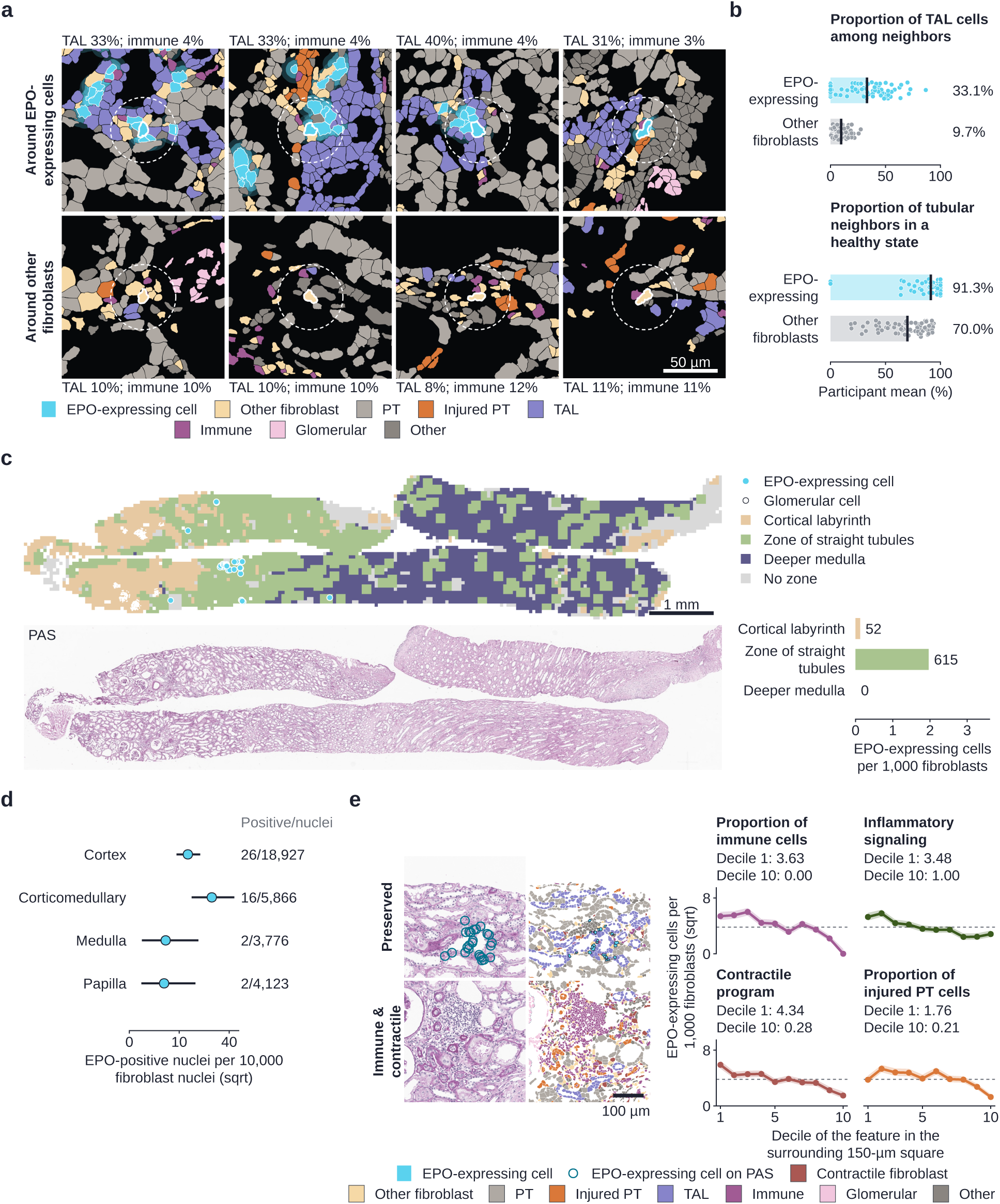
EPO-expressing fibroblasts lie beside intact thick ascending limb in the zone of straight tubules. **a**, The 150-µm neighborhoods of four EPO-expressing cells (top) and of four other fibroblasts (bottom), segmented cells colored by class (key below a); dashed circle, 30 µm; text, proportions of thick ascending limb cells and of immune cells among the 30-µm neighbors; scale bar, 50 µm. **b**, Proportion of thick ascending limb (TAL) cells among the neighbors and proportion of tubular neighbors in a healthy state, for EPO-expressing cells and for other fibroblasts without an *EPO* transcript (points, participant means; bars and numbers, means across participants). **c**, Representative section of the spatial dataset colored by tissue zone (key beside the map), with EPO-expressing cells and glomerular cells, above the same section on PAS, and EPO-expressing cells per 1,000 fibroblasts by zone across all sections (numbers, EPO-expressing cells); scale bar, 1 mm. **d**, EPO-positive fibroblast nuclei per 10,000 fibroblast nuclei of the KPMP single-nucleus dataset by the region of the sample (lines, exact 95% confidence intervals; numbers, EPO-positive over all fibroblast nuclei). **e**, Two 400-µm windows of one section, on PAS (circles, EPO-expressing cells) and as segmented cells (key below e; contractile fibroblasts, contractile program above the upper quartile of all fibroblasts of the spatial dataset): the preserved window holds the most EPO-expressing cells among the windows whose proportions of immune cells and of contractile fibroblasts are both below the medians of the section, and the immune and contractile window has the largest sum of these two proportions, each standardized over the windows of the section; the section is the one with the largest difference in EPO-expressing cells between the two windows. Beside them, EPO-expressing cells per 1,000 fibroblasts by decile of each feature of the surrounding 150-µm square (proportion of immune cells among all cells, inflammatory signaling and contractile program of the fibroblasts, proportion of proximal tubule cells that were injured; bands, 95% confidence intervals; dashed line, overall rate; numbers, rates in the first and tenth deciles); scale bar, 100 µm. PT, proximal tubule.

Of the 684 EPO-expressing cells, 615 were in squares assigned to the zone of straight tubules and none in the deeper medulla (Figure 3c; Supplementary Table S6). Both differences held when the thick ascending limb was left out of the zone assignment (rate ratio within biopsies 2.12; 1.11 to 4.03) and when EPO-expressing cells were compared with fibroblasts of the same zone (43% versus 24%; Supplementary Table S6). *No EPO-expressing cell was found among the 45,716 fibroblasts of the deeper medulla sampled in 97 biopsies, where thick ascending limb cells made up 24% of the cells, against 27% in the zone of straight tubules.* Fibroblasts of the deeper medulla expressed the other genes that characterize EPO-positive fibroblasts more often than fibroblasts of the other zones, but not *EPO* itself (Supplementary Figure S2d). In the KPMP single-nucleus dataset, EPO-positive fibroblast nuclei were likewise most frequent in samples from the corticomedullary region and infrequent in samples from the medulla (Figure 3d).

### EPO-expressing cells are scarce where immune cells are numerous and the contractile program is high

Within the same biopsy, EPO-expressing cells were less frequent in tissue squares with a higher proportion of immune cells (rate ratio per SD 0.20; 0.12 to 0.31), a higher contractile program score of the fibroblasts (0.37; 0.29 to 0.47) and a higher proportion of injured proximal tubule cells (0.55; 0.40 to 0.74; Figure 3e; Supplementary Table S6). The association with immune cells was the strongest and persisted after adjustment for the other features and within the zone of straight tubules. Immune cells of every type were less frequent near EPO-expressing cells (Supplementary Table S4), and EPO-expressing cells were also less frequent where the surrounding fibroblasts had higher inflammatory signaling (0.54; 0.43 to 0.69). EPO-negative fibroblasts with the gene profile (profile-positive cells) were less frequent in squares with more immune cells as well, but less steeply than EPO-expressing cells (rate ratio per SD 0.73; 0.68 to 0.78; Supplementary Table S6).

### EPO-expressing cells are fewer with proximal tubule injury, which accounted for the association with eGFR

Across patients, EPO-expressing cells were fewer at lower eGFR (rate ratio 1.22 per 10 mL/min/1.73 mZ; 1.06 to 1.41; adjusted for diagnosis; Figure 4a; Supplementary Figure S1c,d; Supplementary Table S7; Supplementary Data 4). This was not independent of tubular injury: with the proportion of injured proximal tubule cells in the model, tubular injury was associated with their number (rate ratio per SD 0.40; 0.25 to 0.64) and eGFR was not (1.02; 0.87 to 1.20). In the 104 patients with an eGFR of 60 mL/min/1.73 mZ or higher, EPO-expressing cells were more numerous at lower hemoglobin (rate ratio per g/dL 0.75; 0.58 to 0.95), whereas hemoglobin was not related to their number over all patients (0.81; 0.65 to 1.02).

**Figure 4.**
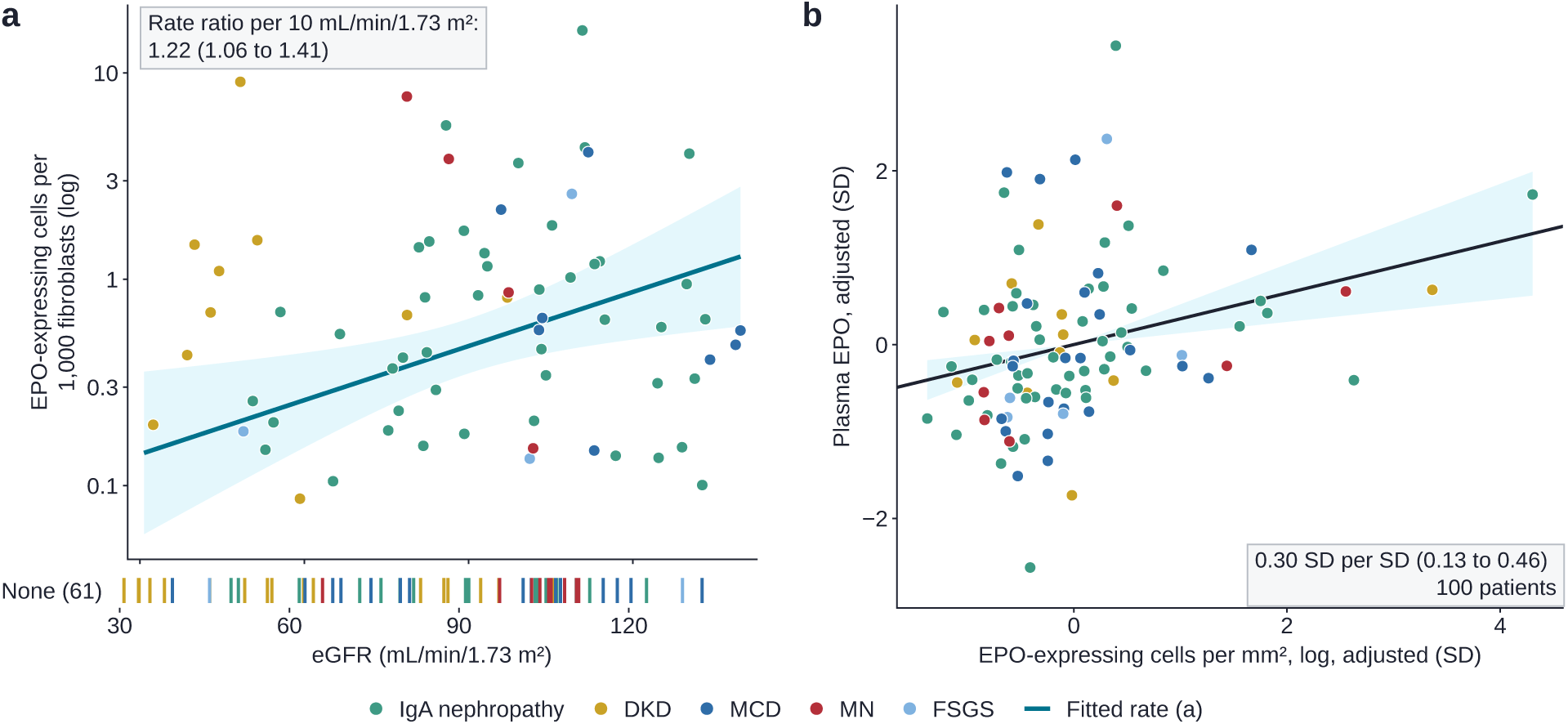
EPO-expressing cells, kidney function and plasma EPO. **a**, EPO-expressing cells per 1,000 fibroblasts plotted against eGFR in the 130 patients (color, diagnosis), with the fitted rate and its 95% confidence band; biopsies without EPO-expressing cells are drawn as ticks below the axis; gray box, rate ratio per 10 mL/min/1.73 m^2^ with 95% confidence interval. **b**, Plasma EPO plotted against the density of EPO-expressing cells in the 100 patients with both measurements, both standardized and adjusted for age, sex, eGFR, assay plate and diagnosis; line and band, the adjusted estimate with its 95% confidence band; gray box, the estimate with 95% confidence interval. DKD, diabetic kidney disease; FSGS, focal segmental glomerulosclerosis; MCD, minimal change disease; MN, membranous nephropathy.

### Plasma EPO is higher in patients with more EPO-expressing cells

Among the 100 patients who had both a biopsy section and a plasma sample collected within 30 days of the biopsy, plasma EPO was higher in those with a higher density of EPO-expressing cells in the biopsy (0.30 SD of plasma EPO per SD of density; 0.13 to 0.46; Figure 4b; Supplementary Table S8).

## Discussion

In kidney biopsies from patients with kidney disease, EPO was expressed by a subset of interstitial fibroblasts with active hypoxia signaling and low TGF-β signaling, located in the zone of straight tubules where the tubules were intact. EPO-expressing fibroblasts were scarce in injured and inflamed interstitium, which may contribute to the insufficient EPO production of kidney disease.

In rodents, EPO-producing fibroblasts express HIF-2α^8^ and lose EPO production when they acquire a myofibroblast phenotype in injured kidneys,^9^ and inflammatory cytokines suppress hypoxia-induced EPO production in cultured cells.^12^ The EPO-positive fibroblasts of the human kidney showed the corresponding features: hypoxia signaling was the most increased and TGF-β signaling the most decreased pathway, and these fibroblasts had lower expression of contractile genes, were scarce among contractile fibroblasts and were almost absent where immune cells were most numerous, whereas fibroblasts with their gene profile but without *EPO* transcripts were only moderately less frequent there. In the few human kidneys previously examined by in situ hybridization, EPO-expressing interstitial cells were detected in only some of the specimens, formed clusters and partly lacked *PDGFRB* transcripts,^14^ as in the present biopsies.

The 684 cells identified here had *EPO* transcripts in tissue fixed at biopsy, and their position could be read against the surrounding segments and cell types. In mice, most fibroblasts of the cortex and outer medulla can produce *EPO* under severe anemia,^30^ whereas cells labeled while producing EPO form a subpopulation that does so repeatedly^11,31^; neither approach shows where such a cell lies in human tissue. In these biopsies the gene profile was present in more fibroblasts than *EPO* itself, in 8,991 profile-positive cells.

Prostacyclin and other prostanoids have been proposed to stimulate EPO production in the kidney through adenylate cyclase and cyclic AMP,^32^ and the genes of EPO-positive fibroblasts included prostacyclin synthase and the phosphodiesterases that degrade cyclic AMP. Coagulation factor VIII and *GREM2*, neither of which has been reported as a marker of EPO-producing cells, were among the genes with the largest differences and were detected more often in EPO-positive fibroblasts in every dataset. These two genes are therefore candidate markers that distinguish EPO-expressing fibroblasts from other fibroblasts in human kidney tissue.

EPO-expressing cells were located in the zone of straight tubules and not in the deeper medulla. In mice, pharmacological prolyl hydroxylase inhibition stabilizes HIF-2α in fibroblasts of the deeper medulla without inducing EPO, whereas deletion of prolyl hydroxylase 2 in fibroblasts induces EPO in all kidney zones^33^; in the present biopsies, profile-positive cells were most frequent in the deeper medulla of all zones (65.6 per 1,000 fibroblasts, against 11.6 in the zone of straight tubules) while no fibroblast there had an EPO transcript, which suggests a similar zonal restriction of EPO expression in the human kidney. TGF-β1 induces methylation and silencing of the *EPO* gene in kidney pericytes,^34^ and EPO-positive fibroblasts had the lowest TGF-β signaling of all pathways, so differences in the accessibility of the *EPO* gene may limit EPO expression in some fibroblasts. Within each zone, EPO-expressing cells differed from other fibroblasts by the same genes and pathway activities, so EPO was expressed by fibroblasts that had both this profile and this location.

EPO-expressing cells were located beside thick ascending limbs, also when compared with fibroblasts of the same zone. Active NaCl reabsorption makes the thick ascending limb one of the largest consumers of oxygen in the kidney, and in the outer medulla, where oxygen supply is limited, this segment is the most vulnerable to hypoxia.^35^ Fibroblasts next to these tubules may experience lower oxygen tension, which would be consistent with their hypoxia signaling, although oxygen tension was not measured. In rats, EPO-producing cells are found mainly in the cortical labyrinth and are recruited from the corticomedullary border outward as hypoxia deepens,^36^ so the zone in which these cells appear may differ between species or with the severity of hypoxia. Thick ascending limb cells were as frequent in the deeper medulla, where EPO was not expressed, so proximity to these tubules alone does not determine EPO expression.

Across patients, EPO-expressing cells were fewer where a higher proportion of proximal tubule cells was injured, in line with the loss of EPO production in injured mouse kidneys^9^ and with the fall in EPO production after severe proximal tubule-specific injury.^37^ Plasma EPO rises as hemoglobin falls while kidney function is preserved,^2^ and EPO-expressing cells were likewise more numerous at lower hemoglobin in patients with an eGFR of 60 mL/min/1.73 mZ or higher. Plasma EPO was higher in patients with more EPO-expressing cells, consistent with a contribution of these cells to circulating EPO. HIF prolyl hydroxylase inhibitors depend in part on kidney cells that retain the capacity to produce EPO^5^; in the present biopsies, EPO-expressing fibroblasts were found mainly where the tubules were intact, in the zone of straight tubules, and whether the extent of this tissue is related to the response to these agents remains to be studied.

This study has several limitations. First, the design was cross-sectional and the biopsies were obtained for clinical indications; most patients had an eGFR of 60 mL/min/1.73 mZ or higher, treatment before the biopsy differed between patients and iron indices were not available, so the associations with eGFR, hemoglobin and plasma EPO apply to early and moderate kidney disease. Second, *EPO* transcripts are sparse: cells defined by two or more transcripts in a single section are those with the highest expression; the whole-transcriptome comparison included 93 nuclei or cells with one or more counts, some of which may represent background; and the larger comparison in tissue sections was limited to the panel genes and may include transcripts from adjacent cells. Third, the findings are based on transcripts in tissue obtained at a single time point; proteins and gene function were not examined, and we could not determine whether fibroblasts in inflamed tissue are lost or cease to express EPO, or whether EPO-expressing fibroblasts are a stable subclass or a reversible state.

## Conclusion

In human kidney disease, EPO is expressed by a subset of interstitial fibroblasts with active hypoxia signaling and low TGF-β signaling, located in the zone of straight tubules adjacent to intact tubules. The fibroblast profile and the proximity to the thick ascending limb were consistent across single-nucleus data, tissue sections and external datasets. This fibroblast state and tissue location are scarce in injured and inflamed tissue. The genes of these cells provide candidate markers for identifying EPO-producing cells in human kidney tissue and indicate signaling pathways for further study.

## Data Sharing Statement

Gene-, cell-type- and model-level summary data underlying the figures and tables are provided in the Supplementary Tables and Supplementary Data 1–4. Individual-level spatial transcriptomic and clinical data are not publicly available because they were generated from biopsy tissue collected under consent for research in Korea. De-identified data can be shared through a collaboration agreement with the KORNERSTONE investigators, subject to approval by the Institutional Review Board and the Biobank of Seoul National University Hospital; requests should be addressed to the corresponding authors. The KPMP single-nucleus and spatial transcriptomics datasets are available from the KPMP data portal (https://atlas.kpmp.org), the external Xenium dataset from Zenodo (https://doi.org/10.5281/zenodo.19868428) and the dataset of PDGFRβ-positive cells from Zenodo (https://doi.org/10.5281/zenodo.4059315). The analysis code is deposited at Zenodo (https://doi.org/10.5281/zenodo.24325319) and is available on request.

## Acknowledgments

The biospecimens and data used for this study were provided by the Biobank of Seoul National University Hospital, a member of the Korea Biobank Network, and by the Korea National Institute of Health (KNIH) research project (project No. KBN5_A03). The results here are in whole or part based upon data generated by the Kidney Precision Medicine Project. Accessed November 8, 2026. https://www.kpmp.org. Funded by the National Institute of Diabetes and Digestive and Kidney Diseases (grant numbers U01DK133081, U01DK133091, U01DK133092, U01DK133093, U01DK133095, U01DK133097, U01DK114866, U01DK114908, U01DK133090, U01DK133113, U01DK133766, U01DK133768, U01DK114907, U01DK114920, U01DK114923, U01DK114933, U24DK114886, UH3DK114926, UH3DK114861, UH3DK114915, UH3DK114937, 1U01DK144965-01 and 1U01DK144994-01).

## Author Contributions

S.G.H. performed the spatial and statistical analyses, prepared the figures and drafted the manuscript. A.K. curated the clinical data, organized and interpreted the results, revised the figures and drafted the manuscript. D.K. performed cell-type annotation of the spatial transcriptomic data; H.K. and H.J.K. generated the spatial transcriptomic data; M.K., Y.S. and Y.-G.K. performed image registration of the spatial data; J.H.J. and J.S.L. generated and processed the single-nucleus RNA-sequencing data; J.-I.O. contributed to the analyses. J.H.K., J.L., M.W.K., H.L., S.S.H., Y.K., J.K., J.H.L., J.P.L., J.I.P., J.T.P., J.N.A. and K.-C.M. contributed to participant enrollment and the collection of samples and clinical and pathology data. S.P. conceived the study, provided the KORNERSTONE samples and clinical data, drafted the manuscript and supervised the analyses. D.K.K. established the cohort and supervised the study. All authors revised the manuscript and approved the final version.

