## Supplemental Information for "Single-cell-level spatial transcriptomics identifies erythropoietin-expressing fibroblasts and their tissue niche in human kidney disease"

**Supplementary Data 1–4 (separate Excel files)**

### Supplementary Methods

#### Participants and clinical data

The 154 participants with a Xenium section of their biopsy formed the spatial cohort. Hemoglobin was taken from the laboratory records of Seoul National University Hospital: the highest value of the day of the biopsy, because several measurements of one day carry no time and hemoglobin falls after a biopsy, or, without a value on that day, the closest earlier value within 90 days (five patients) or the closest later value within 90 days (two patients); for living donors, the highest value of the day of donation. All 154 participants had a value. eGFR at biopsy was calculated with the 2021 CKD-EPI creatinine equation [S1]; values above 150 mL/min/1.73 $m^2$ were checked against the laboratory creatinine of the biopsy day, and one value that the laboratory record did not reproduce was replaced by the eGFR of that creatinine. For living donors, age was the completed age at donation, eGFR the value from pre-donation creatinine and UPCR the pre-donation value of the donor records. Patients were grouped by the diagnosis of the biopsy report; FSGS (5 patients) and membranous nephropathy (13) are reported with their numbers. The participants of every dataset are summarized in Supplementary Table S1 A; clinical values of the external datasets are those of their public releases (KPMP open-access clinical file of 30 September 2026; donor table of the external Xenium dataset), and the deposited data of PDGFR$\beta$-positive cells hold no clinical values.

#### Kidney tissue and histology

Kidney biopsies were fixed in formalin and embedded in paraffin during routine diagnostic processing in the Department of Pathology of Seoul National University Hospital; zero-time biopsies of living donors were obtained at transplantation and processed in the same way. After the Xenium run, the same section was stained with periodic acid–Schiff (PAS) and digitized at 40× magnification (0.263 µm per pixel; Aperio GT450, Leica Biosystems). The PAS images were registered to the cell coordinates of the spatial data by landmark alignment, with a translation for each displayed window fitted to the nuclei of the segmented cells, and registered PAS histology was available for 146 of the 154 participants. PAS images were used to display tissue morphology (Figures 1a, 2b and 3c,e) and did not enter any analysis.

#### Spatial transcriptomics

FFPE sections (5 µm) were mounted on Xenium slides, deparaffinized, decrosslinked, and processed by Xenium Prime hybridization, ligation, rolling-circle amplification, fluorescent labeling, and automated imaging [S2]. The Xenium panel comprised the 5,001-gene Human 5K Pan-Tissue and Pathways base panel plus 100 kidney add-on genes. All sections were acquired with 0.2125-µm in-plane pixels and a 3-µm z-step on Xenium Analyzer software 4.0.1.4 and processed with Xenium Onboard Analysis 4.0.1.0. The Xenium Multi-Tissue Stain informed the instrument's morphology-guided segmentation, which combined stain-defined boundaries, cell interiors, and nuclear expansion. Cell boundaries exported by onboard analysis were used directly; no downstream boundary expansion, imported segmentation, or manual cell resegmentation was applied. Transcripts with Q-score at least 20 were decoded and assigned to segmented cells. The instrument reported a median Q20 decoding fraction of 90.7% (interquartile range, 89.1–92.7%) and a median transcript-assignment fraction of 86.2% (84.4–87.7%) across the 159 sections of the run. Cells with fewer than 10 assigned transcripts and cells without a resolved annotation were excluded, retaining 5,378,790 of 5,991,438 segmented cells (89.8%), of which 5,299,123 carried a label of the joint annotation with the single-nucleus data (Cell annotation). The 154 participants of this study held 155 analyzed sections and 5,150,652 such cells; the cells of the two sections of one participant were combined.

Cells with at least one and with at least two *EPO* transcripts were counted by cell type, and the same counts were taken for each of the 40 negative control probes of the panel. The proportion of fibroblasts with an *EPO* transcript that had two or more was compared with the same proportion for the 40 negative control probes combined by Fisher's exact test. For cell $c$ with counts $x_{cg}$ and library $L_c^0$ without EPO, normalized expression was $y_{cg} = \ln(1 + 100\, x_{cg} / \max(L_c^0, 1))$. With $\mathbf{w}_k$ the loadings of interstitial program $k$ (non-negative matrix factorization programs of fibroblasts, mural and juxtaglomerular cells, unit length over the panel genes, EPO set to zero), the program score of a fibroblast was the cosine projection

$$F_{ck} = \frac{\sum_{g \neq EPO} y_{cg}\, w_{kg}}{\| (y_{cg})_{g \neq EPO} \|_2} \tag{S1}$$

the contractile program is the program in which *ACTA2* has the largest loading and the collagen matrix program the program in which *COL1A1* has the largest loading.

#### Single-nucleus RNA sequencing of the KORNERSTONE cohort

Frozen tissue was minced into pieces smaller than 2 mm and homogenized with a Dounce homogenizer in 2 mL of ice-cold Nuclei EZ lysis buffer (Sigma-Aldrich) supplemented with protease inhibitor (Sigma-Aldrich) and RNase inhibitors (Promega; Thermo Fisher Scientific). After addition of 2 mL of lysis buffer and 5 min on ice, the suspension was passed through a 40-µm strainer (pluriSelect) and centrifuged at 500 × g for 5 min at 4 °C. The pellet was washed in 4 mL of lysis buffer for 5 min on ice, centrifuged again at 500 × g and resuspended in nuclei suspension buffer (1× PBS, 1% bovine serum albumin and 0.1% RNase inhibitor). Nuclei were stained with DAPI and counted with a Countess 3 automated cell counter (Thermo Fisher Scientific). Libraries were prepared with the Chromium Single Cell 3′ Reagent Kit v3 (10x Genomics) according to the manufacturer's protocol and sequenced on an Illumina NovaSeq 6000. Reads were aligned to the GRCh38 pre-mRNA reference (10x Genomics reference GRCh38-2024-A) with Cell Ranger [S3], and the filtered feature–barcode matrices were analyzed in Scanpy (version 1.11.2) [S4]. Nuclei expressing fewer than 300 or more than 9,000 genes or with a mitochondrial transcript fraction of at least 2.5% were removed. Putative doublets were identified with Scrublet [S5] at an expected doublet rate of 7% and removed before integration. The dataset comprised 889,646 nuclei from 125 samples obtained at Seoul National University Hospital and Keimyung University Dongsan Hospital, of which 839,765 nuclei had a cell-type label in the joint annotation with the spatial data described below; their counts of the 5,070 genes shared with the spatial panel were used here.

#### Cell annotation

Cells were annotated following the cell-type hierarchy of the KPMP kidney atlas [S6]. Spatial transcriptomic data were integrated with the single-nucleus RNA-sequencing data from the KORNERSTONE cohort by scVI [S7] and clustered in the shared latent space. Clusters were assigned by canonical markers on the panel, and the single-nucleus profiles of the corresponding clusters were used for markers absent from the panel. Glomerular cells comprised podocytes (*NPHS1*, *NPHS2*, *PTPRQ*), parietal epithelial cells (*CFH*, *ALDH1A2*) and mesangial cells (*ITGA8*, *PIEZO2*, *POSTN*); tubular cells comprised proximal tubule (*LRP2*, *CUBN*, *SLC34A1*), injured proximal tubule (*VCAM1*, *HAVCR1*), loop of Henle (*SLC44A5*, *UMOD*, *SLC12A1*), distal nephron (*SLC12A3*, *CALB1*) and collecting duct cells (*AQP2*, *SLC4A1*, *SLC26A4*); and interstitial cells comprised endothelial cells (*PECAM1*, *EMCN*, *PLVAP*), mural cells (*ACTA2*, *MYH11*), fibroblasts (*DCN*, *C7*, *PDGFRA*), immune cells (*PTPRC*, *CD163*) and Schwann cells (*CDH19*, *SCN7A*). Renin-associated cells were identified by *REN* expression, and adipocytes and proliferating cells formed classes of their own. Every spatial cell took its label from this joint annotation, the same annotation that classed the single nuclei; annotation did not use diagnosis, and the same labels were used for all analyses in this study. The thick ascending limb and its adaptive state and the injured proximal tubule were labels of the joint annotation; injured proximal tubule cells were divided further by their own *VCAM1* and *HAVCR1* transcripts and immune cells by the marker-defined subtypes used previously for these data, the S1 or S2 and the S3 segments of the proximal tubule were assigned within the proximal tubule labels by the segment definitions of the KPMP kidney atlas

[S6], as applied to these data previously [S8], and endothelial cells outside glomeruli were kept as one class.

#### EPO-expressing cells compared with matched fibroblasts

For each EPO-expressing cell, ten fibroblasts of the same participant without an *EPO* transcript were drawn at random, without reuse, from the 30 with the nearest log library size. Each gene detected in at least 1% of these cells (2,743 genes, EPO excluded) was compared by

$$\Pr(D_{cg} = 1) = 1 - \exp\left[-\exp\left(\beta_{0g} + \beta_g \, EPO_c + \alpha_{g,p(c)} + \ln\max(L_c, 1)\right)\right] \tag{S2}$$

a binomial model with complementary log–log link, participant fixed effects $\alpha_{g,p}$ and log library size as offset; $\exp(\beta_g)$ is the rate ratio of detection. Because cells of one participant are not independent, standard errors were clustered by participant, and genes detected more or less often were those with $q < 0.05$ (Benjamini–Hochberg [S9]) with these errors. The *z* statistics of the model were compared between the members of each Hallmark set [S10] with at least ten tested genes and the other genes by the Mann–Whitney test, with Benjamini–Hochberg *q* over the sets. The proportion of EPO-expressing and of matched fibroblasts with at least one of the transcripts of *F8*, *GREM2*, *PTGIS* and *VEGFA* was counted without a test. In a second comparison, the ten fibroblasts of each EPO-expressing cell were drawn in the same way from the fibroblasts of the same participant that were located in the same tissue zone (the zone of the 50-µm square holding the cell; see Neighborhoods, tissue squares and zones), all of them when the zone held fewer than ten; the 17 EPO-expressing cells in squares without a zone were left out, and the tested genes, the model, the clustered errors, the *q* values and the pathway activity were those of the first comparison.

EPO-expressing cells of a section within 30 µm of each other were joined into groups (single linkage: the connected components of the graph whose edges join cells within 30 µm), and the proportion of these cells with another EPO-expressing cell within 30 µm was compared with the proportion expected when the same number of fibroblasts of each section was labeled at random (1,999 relabelings).

#### EPO transcripts in other cell types

For each cell other than a fibroblast with two or more *EPO* transcripts, the distance between its boundary and that of the nearest EPO-expressing fibroblast was measured and compared with that of ten matched cells of the same type from the same biopsy without an *EPO* transcript and with a similar library size; cells within 5 µm were considered in contact, and contact was compared by logistic regression with standard errors clustered by participant. The proportion of *EPO* transcripts located within the nucleus was compared between these cells and EPO-expressing fibroblasts.

#### EPO-positive cells by cell class

In each single-cell dataset, the cells or nuclei with one or more and with two or more *EPO* counts were counted by cell class (Supplementary Table S1; Figure 1c–e): in the single-nucleus data from the KORNERSTONE cohort, all 839,765 nuclei by the classes of the joint annotation; in the KPMP single-nucleus dataset [S6] (release of 18 May 2025; 615,169 nuclei of 157 participants), all nuclei by the first-level subclasses of its annotation grouped into the same classes; in the dataset of PDGFR$\beta$-positive cells [S11], by the authors' annotation; and in the external Xenium dataset, by the authors' annotation grouped into these classes. In the two single-nucleus datasets, the frequency of nuclei with one or more (and with two or more) *EPO* counts among fibroblast nuclei and among all other nuclei was compared by the rate ratio, with an exact 95% interval conditional on the total number of such nuclei (the Clopper–Pearson interval of the fibroblast proportion of these nuclei, transformed to the rate ratio). In KPMP, nuclei with an *EPO* count were also counted in each subclass of the fibroblast and of the vascular smooth muscle and pericyte classes, and fibroblast nuclei with an *EPO* count were counted by the region of the sample (cortex, corticomedullary region, medulla and papilla), taken from the region field of the cell metadata of the release, with exact (Clopper–Pearson) 95% intervals of the proportion.

#### Single nuclei over the whole transcriptome

For every panel gene, the mean expression of each cell type of the spatial dataset was computed over all cells (pooled counts per $10^4$); a gene whose highest cell type was not the fibroblast but one of the cell types common within 30 µm of EPO-expressing cells (thick ascending limb, distal convoluted tubule, connecting tubule, principal and intercalated cells, proximal tubule and extraglomerular endothelium) was marked as an adjacent-cell transcript.

EPO-positive fibroblast nuclei were compared with matched fibroblast nuclei of the same participant (ten drawn from the 30 with the nearest log library size) by equation (S2) in three datasets: the KPMP single-nucleus dataset (45 nuclei of 29 participants, whole transcriptome; the 46th nucleus had no fibroblast nucleus without an *EPO* count left in its participant), the single-nucleus data from the KORNERSTONE cohort (22 nuclei of 14 samples, the 5,070 genes shared with the spatial panel) and the dataset of PDGFR$\beta$-positive cells (Zenodo, https://doi.org/10.5281/zenodo.4059315, file Human_PDGFRb.tar.gz), in which the cells annotated by the authors as fibroblasts, myofibroblasts or pericytes with an *EPO* count (26 cells of 6 patients) were compared with ten such cells of the same patient. Genes detected in at least 1% of the cells of a dataset were tested, and separated or non-converged fits were left out. For each gene, the log rate ratios $b_d$ with standard errors $s_d$ of the datasets *d* in which it was tested were combined as

$$\widehat{\beta} = \frac{\sum_d b_d/s_d^2}{\sum_d 1/s_d^2}, \qquad SE(\widehat{\beta}) = (\sum_d 1/s_d^2)^{-1/2} \tag{S3}$$

with Benjamini–Hochberg *q* over the 16,217 genes tested in any dataset.

The genes detected more often were tested for over-representation of Gene Ontology biological processes [S12, S13] (annotations propagated to ancestor terms), of Reactome pathways [S14] and of imprinted genes (the human list of Geneimprint [S15], accessed 14 November 2026, with the status imprinted and predicted kept apart), by the hypergeometric test with all tested genes as background, with Benjamini–Hochberg *q* within each collection. Sets at $q < 0.05$ were grouped by their genes: sets of more than 1,000 tested genes were left out as broad, and sets that shared their genes were joined into one group. The combined *z* of all tested genes was compared between the members of each Hallmark set with at least ten tested genes and the other genes by the Mann–Whitney test, with Benjamini–Hochberg *q* over the sets. The 70 genes detected more often were also checked against the Hallmark hypoxia set, the Reactome sets on hypoxia, the Gene Ontology process response to hypoxia with its descendants and the Norn cell markers of EPO-producing cells of the mouse [S16] mapped to human genes, and the rows of the combined comparison are given for eleven genes tied to EPO-producing cells in earlier studies [S16–S20].

Transcription factor activity was inferred from the combined comparison by a univariate linear model [S21]: for factor *f* with signed CollecTRI weights $m_{fg} \in \{-1, 0, 1\}$ over the genes *g* [S22],

$$z_g = a_f + b_f\, m_{fg} + \varepsilon_g \tag{S4}$$

fitted by least squares over all tested genes; the activity is the *t* statistic of $b_f$, for factors with at least five targets among the genes, with Benjamini–Hochberg *q* over the factors. The activity of the 14 signaling pathways of the human PROGENy model [S23] was inferred by the same model with the weights of the 500 response genes of each pathway with the smallest *P* in the model (the other genes weighted 0) in place of $m_{fg}$, with *q* over the 14 pathways. The contribution of a response gene to the activity of the hypoxia or the TGF-β pathway was its weight times its combined *z*; Figure 2d names the five genes with the largest positive contribution to the hypoxia activity and the five with the largest negative contribution to the TGF-β activity, which was negative, and the 20 response genes with the largest contribution in the direction of each activity are listed in Supplementary Table S3 D (all response genes in Supplementary Data 2). For tissue sections, the same pathway model was fitted to the *z* statistics of the comparison of EPO-expressing with matched fibroblasts, each the log rate ratio of equation (S2) over its standard error clustered by participant, over the 2,743 tested panel genes; the response genes of each pathway on the panel are given with the activities (Supplementary Table S3).

#### Profile-positive cells

The profile score of a fibroblast was the mean of $y_{cg}$ over the genes detected more often in EPO-positive fibroblast nuclei with the datasets combined that were on the panel and were not adjacent-cell transcripts (EPO excluded). Fibroblasts without an *EPO* transcript whose score reached the median score of the EPO-expressing cells were called profile-positive cells. Profile-positive cells were also counted per 1,000 fibroblasts of each tissue zone, by the zone of the square holding the cell, and in the squares and deciles of the proportion of immune cells used for EPO-expressing cells (Wilson intervals); their number per square was related to the proportion of immune cells by the model of equation (S5) over the squares of the participants with a profile-positive cell.

Fibroblast nuclei of the KPMP single-nucleus dataset were scored as $\frac{1}{|G|}\sum_{g\in G}\ln(1+10^4\,x_g/L^0)$ over the genes *G* detected more often in EPO-positive nuclei at $q < 0.05$ when only the other two datasets were combined (20 genes), with $L^0$ the counts of the nucleus without EPO, and the percentile of each EPO-positive nucleus among the nuclei without an *EPO* count was taken. The 32,863 fibroblast nuclei of KPMP were embedded by the usual preprocessing without clustering: counts per $10^4$ and log transformation, 2,000 variable genes by normalized dispersion within 20 bins of mean expression, scaling clipped at 10, 30 principal components and UMAP [S24] with 15 neighbors and a minimum distance of 0.5.

#### Neighborhoods, tissue squares and zones

Within each section, the distance from each fibroblast to the nearest cell of each kind was found for kinds with at least three cells in the section, and the neighborhood of a cell was the 20 nearest cells within 30 µm. The proportions of cell types among the neighbors and the distances were averaged per participant for EPO-expressing and for other fibroblasts and compared by the Wilcoxon signed-rank test, with Benjamini–Hochberg *q* within the distances and within the neighbor proportions.

The tissue was divided into 50-µm squares *b*. For each square, the proportion of proximal tubule cells that were injured (proportion of injured proximal tubule cells), the proportion of immune cells among all cells and the mean collagen matrix and contractile program scores of the fibroblasts other than EPO-expressing cells were taken over the $3 \times 3$ block of squares around it (150 µm), and $P_b$ was the number of proximal tubule cells of the block. Squares with $P_b \geq 5$ in the 77 participants with an EPO-expressing cell in such squares entered

$$\log E[Y_b] = \log S_b + \beta_0 + \beta^\top \mathbf{z}_b + \gamma \log(1 + P_b) + \alpha_{p(b)} \tag{S5}$$

a Poisson model of the EPO-expressing cells $Y_b$ with the fibroblasts $S_b$ as exposure, the features $\mathbf{z}_b$ standardized over these squares, participant fixed effects and standard errors clustered by participant, because the features of adjacent squares overlap; each feature was fitted alone, and the proportion of immune cells, the contractile program, the proportion of injured proximal tubule cells and the collagen matrix program were also fitted together. For the inflammatory signaling of fibroblasts, each fibroblast *c* was scored for the TNF-α, NF-κB and JAK–STAT pathways *k* of the human PROGENy model [S23] as $s_{ck} = \sum_g w_{kg}\, y_{cg}$ over the response genes of the pathway on the panel (the 500 genes with the smallest *P* of each pathway's model; 214, 227 and 181 genes on the panel), and

$$I_c = \frac{1}{3}\sum_k \frac{s_{ck} - \bar{s}_k}{SD(s_k)} \tag{S6}$$

with mean and standard deviation over all 826,184 fibroblasts; the feature of a square was the mean of $I_c$ over the fibroblasts other than EPO-expressing cells of its $3 \times 3$ block, as for the program scores, and the three pathway scores entered the same models one at a time. The single-feature models were also fitted over the squares of the zone of straight tubules alone (below), with the features standardized over all modeled squares.

For the tissue zones, every square holding a fibroblast (276,824 squares) was placed by the cells of its $3 \times 3$ block. Convoluted segments were glomerular cells (podocytes, parietal epithelial and mesangial cells), proximal tubule of the S1 and S2 segments (injured proximal tubule by its segment), distal convoluted and connecting tubule; straight segments were proximal tubule of the S3 segment, thick ascending limb (with its adaptive state), descending thin limb and collecting duct (principal and intercalated cells). A square was placed in the cortical labyrinth when its block held more convoluted than straight cells, in the zone of straight tubules (medullary rays and outer stripe) when the block held more straight than convoluted cells and a proximal tubule cell, and in the deeper medulla (inner stripe and below) when the block held more straight than convoluted cells, a thin limb cell and no proximal tubule cell; other squares had no zone. EPO-expressing cells were counted per 1,000 fibroblasts of each zone, and the proportion of thick ascending limb cells (with the adaptive state) was counted among all cells of the squares of each zone. Zones were compared within biopsies by a Poisson model with the zone, participant fixed effects, the fibroblasts of the square as exposure and standard errors clustered by participant, over the squares with a zone of the 80 participants with an EPO-expressing cell. The zones were assigned a second time with the thick ascending limb and its adaptive state left out of the straight segments, which then comprised proximal tubule S3, thin limb and collecting duct, and compared in the same way. Within each zone, the proportion of thick ascending limb cells among the 20 nearest cells within 30 µm was averaged per participant for EPO-expressing cells and for fibroblasts without an *EPO* transcript and compared by the Wilcoxon signed-rank test over the participants with an EPO-expressing cell in the zone. Clustering was assessed by reassigning EPO expression at random among the fibroblasts of the same biopsy and zone 1,000 times, keeping the number of EPO-expressing cells; the proportion of EPO-expressing cells with another EPO-expressing cell within 30 µm was compared with its distribution under reassignment, with the 17 EPO-expressing cells in squares without a zone left out.

#### Cells adjacent to EPO-expressing cells

The 30-µm neighbors of EPO-expressing cells and of their matched fibroblasts were described by the segment and state labels of the annotation. For each measure, the participant means of the neighbors of EPO-expressing and of matched fibroblasts were compared by the Wilcoxon signed-rank test with Benjamini–Hochberg *q* over the measures.

Thick ascending limb cells in a healthy state within 30 µm of an EPO-expressing cell were compared with those within 30 µm of zone-matched fibroblasts from the same participant; cells adjacent to both were excluded, and EPO-expressing and matched fibroblasts without such a cell within 30 µm did not enter. Detection of *UMOD*, *KCNJ1* and *ATP1A1* (*SLC12A1* and *ATP1B1* are not on the panel), the Hallmark oxidative phosphorylation score calculated from the panel genes and the activity of the hypoxia pathway were compared by paired Wilcoxon signed-rank tests of participant means. The Benjamini–Hochberg procedure was applied within each analysis.

#### Participant-level models

EPO-expressing cells $Y_i$ of participant *i* were modeled by negative binomial regression,

$$\log \mu_i = \log S_i + \beta_0 + \beta_1 x_i + \gamma^\top (age_i, female_i, e_i, T_i), \qquad Var(Y_i) = \mu_i + \alpha \mu_i^2 \tag{S7}$$

with $S_i$ the fibroblasts of the section, $x_i$ the exposure (eGFR $e_i$ per 10 mL/min/1.73 m$^2$, hemoglobin per g/dL or diagnosis), $T_i$ the proportion of thick ascending limb cells among the section's cells and $\alpha$ estimated by maximum likelihood; the eGFR model was fitted over all patients with diagnosis added and separately in IgA nephropathy and in DKD, the two largest diagnoses. The proportion of injured proximal tubule cells of the section (per SD) was added to the eGFR model over all patients. The dispersion was estimated by maximizing the profile likelihood, and coefficients and Wald intervals came from the negative binomial model at that dispersion. Hemoglobin was related to EPO-expressing cells in all 130 patients and in those with an eGFR of 60 mL/min/1.73 m$^2$ or more and below 60.

#### Plasma proteomics and plasma EPO

Plasma was separated from blood collected in EDTA tubes and stored for the KORNERSTONE cohort until analysis. Plasma proteins were measured with the Olink Explore HT platform, a proximity extension assay in which each protein is bound by a pair of oligonucleotide-labeled antibodies and the resulting DNA reporter is quantified by next-generation sequencing [S25]. Protein levels are expressed as normalized protein expression (NPX) on a $\log_2$ scale after the quality control and plate normalization of the manufacturer. For each patient with a spatial section, the plasma sample nearest the biopsy within 30 days was used (100 patients on six plates; median 0 days from the biopsy), and plate entered every model as a categorical covariate. Plasma EPO (Olink assay OID43684, NPX) was related to the density of EPO-expressing cells, $\ln(1 + \text{cells per mm}^2)$ standardized over the patients, with age, sex, eGFR, plate and diagnosis, both as NPX and standardized, by least squares with HC3 errors [S26].

#### External spatial datasets

The KPMP spatial transcriptomics dataset (Visium; 210 kidney samples of 209 participants) was read from the open-access data release. Each tissue spot was scored for signatures as the mean of $\log(1 + \text{counts per } 10^4)$ over its genes (thick ascending limb, *UMOD*, *SLC12A1*, *CLDN16* and *EGF*; proximal tubule, *LRP2*, *CUBN*, *SLC34A1* and *SLC5A12*; injured proximal tubule, *HAVCR1*, *VCAM1*, *PROM1* and *CDH6*; and immune cells, *PTPRC*, *CD74*, *LYZ* and *CD3E*), standardized within each sample. Each spot with at least one *EPO* UMI was compared with the ten spots without *EPO* of the same sample nearest in log total UMI,

$$\Delta_k = \frac{1}{|P|} \sum_{j \in P} \left( x_{jk} - \frac{1}{10} \sum_{m \in C_j} x_{mk} \right) \tag{S8}$$

with *P* the *EPO*-positive spots, $C_j$ the matched spots of spot *j* and $x_{jk}$ the standardized score of signature *k*; intervals came from 2,000 resamples of participants, and the direction per sample was tested by the sign test. The same difference was computed with the mean score of the six spots adjacent to each spot (spots within 1.1 times the center-to-center distance of the array) in place of the score of the spot itself, for the EPO-positive and the matched spots alike.

The external Xenium dataset [S27] (16 participants: 8 with DKD, 5 with other kidney diseases and 3 controls; 951,040 cells) was read from its public deposit (Zenodo, https://doi.org/10.5281/zenodo.19868428) with its authors' cell annotations, grouped into the cell classes used here, and fibroblasts with two or more *EPO* counts were compared with matched fibroblasts as in the spatial dataset. The proportion of thick ascending limb cells among the 20 nearest cells within 30 µm was averaged per sample for these fibroblasts and for fibroblasts without an *EPO* count, without a test.

#### Software

Analyses used Python 3.14 (pandas, NumPy, SciPy, statsmodels, scikit-learn, networkx, umap-learn, shapely 2.2) and R 4.6 (ggplot2, patchwork, ggrepel).

### Supplementary Results

#### *EPO* transcripts by cell type

Cells with one or more *EPO* transcripts occurred at 11 to 31 per 10,000 cells across cell types, and no negative control probe reached two transcripts in more than two fibroblasts (Supplementary Table S1). Of the 1,031 cells with two or more *EPO* transcripts, 684 were fibroblasts of 81 participants, followed by thick ascending limb (130), endothelial (86) and immune cells (45). Of these 347 other cells, 163 (47%) lay within 5 µm of an EPO-expressing fibroblast, compared with 13 of 3,470 matched cells of the same types (0.4%; Supplementary Table S1). In the KPMP single-nucleus dataset, nuclei with two or more *EPO* counts numbered 6.4 per 10,000 fibroblast nuclei, compared with 1.1 in the vascular smooth muscle and pericyte class. EPO-expressing cells were found in 12 of the 24 living donors, 44 of the 60 patients with IgA nephropathy and 10 of the 28 with DKD (Table 1), and in 56 of the 104 patients with an eGFR of 60 mL/min/1.73 $m^2$ or more and 13 of the 26 with a lower eGFR. Nearly two thirds of the EPO-expressing cells (65%) had another EPO-expressing cell within 30 µm, 27 times the proportion expected for fibroblasts of the same section (Supplementary Data 1) and 17.5 times that expected for fibroblasts of the same section and zone (Supplementary Table S1). Of the 684 EPO-expressing cells, 239 had no other EPO-expressing cell within 30 µm and 445 formed 119 groups of two or more cells (median 2 cells, largest 26), found in 46 of the 81 biopsies with EPO-expressing cells. EPO-expressing cells by diagnosis, eGFR and anemia are shown in Supplementary Figure S1c,d.

#### Genes of EPO-expressing cells

In tissue sections, 101 genes were detected more often and 124 less often in EPO-expressing cells than in matched fibroblasts ($q < 0.05$; Supplementary Table S2). Of the 70 genes of the nuclei, 34 were on the panel and 29 were detected in at least 1% of the cells; 21 of the 29 were detected more often in EPO-expressing cells, among them *F8* (15% versus 0.3% of cells), *PTGIS* (32% versus 2%), *VEGFA* (25% versus 3%) and *GREM2* (13% versus 0.7%), and the eight others (*ABCC9*, *APLF*, *CCDC137*, *CCL11*, *DNPEP*, *F2R*, *FHL1* and *SRCAP*) did not differ (Supplementary Figure S2b). Genes with their highest expression in tubular cells, among them *UMOD* (36% versus 13% of cells), were detected more often in EPO-expressing cells in tissue sections and not in EPO-positive nuclei, and genes of oxidative phosphorylation, which were lower in EPO-positive nuclei, ranked higher in tissue sections (*q* 0.02); these transcripts are abundant in the adjacent thick ascending limb. Targets of MYC were also detected less often in EPO-positive nuclei, which together with the lower oxidative phosphorylation and the imprinted genes is compatible with quiescent cells with low biosynthetic activity. *CXCL12* and *PDGFRA* were detected less often in EPO-expressing cells (17% versus 34% and 16% versus 25% of cells). *PDGFRB* was detected in 12% of EPO-expressing cells, compared with 22% of matched fibroblasts, and 56% of EPO-expressing cells contained at least one transcript of *F8*, *GREM2*, *PTGIS* or *VEGFA*, compared with 6% of matched fibroblasts. In tissue sections, the Hallmark epithelial–mesenchymal transition set was the lowest-ranked set (*q* $2.0 \times 10^{-8}$; Supplementary Table S2).

In the three single-cell datasets, the 70 genes were detected more often in 93 EPO-positive fibroblast-lineage nuclei and cells of 49 participants or samples than in matched nuclei and cells of the same participants. In KPMP, *VEGFA* was detected in 53% of EPO-positive versus 9% of other fibroblast nuclei, *GREM2* in 36% versus 4%, *PTGIS* in 49% versus 15% and *PDE8B* in 62% versus 21%. The endothelial genes *PECAM1*, *EMCN* and *KDR* were not detected more often in EPO-positive nuclei (*P* 0.088, 0.959 and 0.071), and in KPMP the endothelial gene *EGFL7* was detected in 4% of EPO-positive nuclei, compared with 42% for *F8*. Imprinted genes (*H19*, *IGF2*, *MEG3* and *MEG8*) were 13 times as frequent among the 70 genes as expected ($P = 2.4 \times 10^{-4}$). Signaling by G protein-coupled receptors was enriched (*q* 0.003), with receptors for endothelin (*EDNRB*), thrombin (*F2R*), relaxin (*RXFP1*) and vasoactive intestinal peptide (*VIPR2*), as was the metabolism of cyclic nucleotides (*q* 0.007). Collagen formation was enriched (*q* 0.008), with the collagen-modifying enzymes *P4HA1*, *PLOD2* and *LOXL3*, and so was the regulation of body fluid levels ($q$ 0.014), with *F8*, *VEGFA*, *EDNRB*, *F2R* and the natriuretic peptide receptor *NPR3*. Two smaller groups, leukocyte chemotaxis and muscle cell development, were also enriched ($q$ 0.026 to 0.049; Supplementary Table S3). The Hallmark set of hypoxia-responsive genes as a whole was not higher ($P = 0.13$), although *VEGFA*, *P4HA1* and *TGFBI* of this set were among the 70 genes. *EPAS1* was detected more often in EPO-positive nuclei ($P < 0.001$) and *PDGFRB* less often ($P = 0.014$), and of 644 transcription factors with at least five targets among the genes, 24 had an inferred activity at $q < 0.05$, 15 of them higher in EPO-positive nuclei, among them EPAS1, HIF1A and their partner ARNT ($q$ 0.036 to 0.046; Supplementary Table S3). Of the 70 genes, three were members of the Hallmark hypoxia set (*VEGFA*, *TGFBI* and *P4HA1*), *VEGFA* of the two Reactome sets on hypoxia, three of the Gene Ontology process response to hypoxia (*VEGFA*, *PTGIS* and *PLOD2*) and three of the Norn cell markers of the mouse (*ITIH5*, *F2R* and *COL6A1*); *F8* and *GREM2* were in none of these lists (Supplementary Table S3). In tissue sections, the activity of the hypoxia pathway was the highest and that of the TGF-β pathway the lowest of the 14 pathways (Supplementary Table S3). With comparators taken from the same tissue zone of the same biopsy, 20 of the 21 genes remained detected more often in EPO-expressing cells; the activity of the hypoxia pathway was 7.57 ( $q < 0.001$) and that of the TGF-β pathway $-3.24$ ($q$ 0.008) (Supplementary Tables S2 and S3).

In the external Xenium dataset, 172 of the 241 cells with two or more *EPO* counts, found in 13 of 16 participants, were fibroblasts (11 participants), and the matched comparison agreed in sign with the spatial dataset of this study for 163 of the 194 shared genes at $q < 0.05$ here and for all 41 genes significant in both (Spearman $\rho$ 0.30 over 2,407 shared genes; Supplementary Figure S2c).

#### Fibroblast subclasses

In KPMP, EPO-positive nuclei were found in every fibroblast subclass except one, most often in the main fibroblast subclass (29 of 46 nuclei), and 1 of the 4,071 myofibroblast nuclei contained an *EPO* count; in the dataset of PDGFR$\beta$-positive cells, *EPO* counts were found in 12 of 13,130 myofibroblasts and 10 of 6,964 fibroblasts (Supplementary Figure S1b; Supplementary Table S1).

#### Profile-positive cells

Scored on the 20 genes found in the other two datasets alone, 2,078 KPMP fibroblast nuclei (6.3%) expressed the profile without an *EPO* count, and the EPO-positive nuclei ranked at a median 94th percentile of the score of the nuclei without an *EPO* count (Supplementary Table S3). Nuclei of the medullary fibroblast subclass without an *EPO* count also scored high on the profile (median 88th percentile). In the spatial dataset, 8,991 fibroblasts without an *EPO* transcript (1.1% of the 826,184 fibroblasts) expressed the gene profile of EPO-positive fibroblasts; they were present in 153 of the 154 biopsies (Supplementary Table S3). Profile-positive cells numbered 4.1, 11.6 and 65.6 per 1,000 fibroblasts in the cortical labyrinth, the zone of straight tubules and the deeper medulla (Supplementary Figure S2d; Supplementary Table S3). In the first and the tenth decile of the proportion of immune cells, profile-positive cells numbered 7.0 and 2.2 per 1,000 fibroblasts, and EPO-expressing cells 3.6 in the first decile and none in the tenth (Supplementary Table S6).

#### Location

The median distance from an EPO-expressing cell to the nearest thick ascending limb cell was 12.0 µm and from other fibroblasts 47.2 µm (difference of participant medians $-24.8$ µm; $-38.4$ to $-19.8$; lower in 72 of 81 participants); 86% versus 28% were located within 20 µm of a thick ascending limb cell. Proximal tubule cells were located nearer to EPO-expressing cells than to other fibroblasts (median difference $-17.4$ µm), injured proximal tubule did not differ, and immune cells were located farther (9.5 µm). EPO-expressing cells were located nearer to endothelial cells than other fibroblasts were (18 µm versus 22 µm) and had fewer vascular smooth muscle cells (3% versus 6% of the neighbors) and fewer immune cells (4% versus 11%) nearby (Supplementary Figure S3a,b; Supplementary Table S4). Adjacent to EPO-expressing cells, fewer thick ascending limb cells were in an adaptive state (8% versus

38%) and fewer proximal tubule cells in an injured state (11% versus 31%), and proximal tubule neighbors were more often of the S3 segment (36% versus 22%; Supplementary Figure S3c; Supplementary Table S5). KPMP spatial transcriptomics spots with *EPO* transcripts had a higher thick ascending limb score than depth-matched spots of the same sample (0.36 SD; 0.27 to 0.48), and in the external Xenium dataset, thick ascending limb cells made up 21% of the neighbors of fibroblasts with two or more *EPO* counts, compared with 5% of those of other fibroblasts (means of 11 samples; Supplementary Figure S3d). Healthy thick ascending limb cells adjacent to EPO-expressing cells did not differ from those adjacent to zone-matched fibroblasts in the detection of *UMOD*, *KCNJ1* and *ATP1A1*, the oxidative phosphorylation score or the activity of the hypoxia pathway (Supplementary Table S5).

EPO-expressing cells numbered 0.12 and 1.96 per 1,000 fibroblasts in the cortical labyrinth and the zone of straight tubules, with a within-biopsy rate ratio of 11.6 (7.1 to 19.1), and none were found in the deeper medulla (Supplementary Table S6). With the thick ascending limb left out of the zone assignment, 362 of the 684 EPO-expressing cells were in the zone of straight tubules (1.83 per 1,000 fibroblasts, against 0.56 in the cortical labyrinth) and none in the deeper medulla. Within the zone of straight tubules, thick ascending limb cells were more frequent among the neighbors of EPO-expressing cells than among those of other fibroblasts in 62 of 71 participants, and also within both zones of the second assignment (Supplementary Table S6). In KPMP, EPO-positive fibroblast nuclei numbered 27 per 10,000 fibroblast nuclei in samples of the corticomedullary region, compared with 14 in cortical and 5 in medullary samples. Within biopsies, EPO-expressing cells were unrelated to the collagen matrix program of the fibroblasts (rate ratio per SD 1.06; 0.59 to 1.90; Supplementary Table S6).

#### EPO-expressing cells and patients

In the two largest diagnoses, the rate ratio per 10 mL/min/1.73 m² was 1.23 (1.01 to 1.51) in IgA nephropathy (60 patients) and 0.93 (0.66 to 1.33) in DKD (28 patients). With the proportion of injured proximal tubule cells in the model, the rate ratio for eGFR over all patients was 1.02 (0.87 to 1.20) (Supplementary Table S7).

#### Supplementary Tables

**Supplementary Table S1.** Datasets and EPO-positive cells by cell type

*A. Participants of each dataset*

| Dataset | Participants | Samples or sections | Composition | Age, years | Female, *n* (%) | eGFR, mL/min/1.73 m$^2$ | Cells or nuclei | Fibroblasts | EPO-positive (participants) |
|---|---|---|---|---|---|---|---|---|---|
| Spatial dataset | 154 | 155 | IgAN 60; DKD 28; living donors 24; MCD 24; MN 13; FSGS 5 | 48 (35, 57) | 72 (47%) | 98 (76, 107) | 5,150,652 | 826,184 | 684 (81) |
| Single-nucleus data from the KORNERSTONE cohort | 125 | 125 | IgAN 36; MN 27; FSGS 22; MCD 16; nephrectomy controls 15; DKD 9 | 48 (35, 62) | 49 (39%) | 93 (74, 106) | 839,765 | 25,297 | 22 (14) |
| KPMP single-nucleus dataset | 157 | – | CKD 72; healthy reference 37; AKI 33; DM-R 11; not in the clinical file 4 | 50–59 (band) | 70 (46%) | 67 (47, 95) | 615,169 | 32,863 | 46 (29) |
| Dataset of PDGFR$\beta$-positive cells | 8 | – | mesenchymal 25,385; endothelial 2,341; immune 2,083; epithelial 461 | – | – | – | 30,270 | 23,883 | 26 (6) |
| KPMP spatial transcriptomics dataset (Visium) | 209 | 210 | CKD 95; AKI 62; healthy reference 28; DM-R 24 | 50–59 (band) | 95 (45%) | 75 (48, 97) | 180,176 | – | 1,724 (137) |
| External Xenium dataset | 16 | 16 | DKD 8; controls 3; AA amyloidosis 2; IgAN 1; MN 1; C3GN 1 | 64 (48, 72) | 5 (31%) | – | 951,040 | 120,457 | 172 (11) |
| Plasma (Olink Explore HT) | 100 | 100 | IgAN 52; MCD 23; DKD 11; MN 9; FSGS 5 | 42 (33, 54) | 44 (44%) | 97 (78, 111) | – | – | – |

*B. EPO-positive cells or nuclei by cell class*

| | KORNERSTONE (nuclei) | | | | KPMP (nuclei) | | | | External Xenium dataset (cells) | | |
|---|---|---|---|---|---|---|---|---|---|---|---|
| Cell class | Nuclei | *EPO* ≥1 | ≥1 per 10,000 | *EPO* ≥2 | Nuclei | *EPO* ≥1 | ≥1 per 10,000 | *EPO* ≥2 | Cells | *EPO* ≥2 | ≥2 per 10,000 |
| Glomerular epithelium | 12,788 | 6 | 4.7 | 0 | 15,600 | 4 | 2.6 | 0 | 35,509 | 0 | 0.0 |
| Proximal tubule | 216,705 | 69 | 3.2 | 0 | 178,531 | 64 | 3.6 | 4 | 296,098 | 3 | 0.1 |
| Thin limb | 39,460 | 3 | 0.8 | 0 | 24,916 | 7 | 2.8 | 1 | 10,822 | 0 | 0.0 |
| Thick ascending limb | 246,033 | 67 | 2.7 | 2 | 140,842 | 53 | 3.8 | 0 | 113,932 | 3 | 0.3 |
| Distal convoluted and connecting tubule | 91,782 | 20 | 2.2 | 0 | 50,160 | 15 | 3.0 | 0 | 58,869 | 0 | 0.0 |
| Collecting duct | 113,888 | 30 | 2.6 | 0 | 84,015 | 35 | 4.2 | 2 | 103,682 | 4 | 0.4 |
| Endothelium | 61,225 | 19 | 3.1 | 0 | 34,616 | 5 | 1.4 | 0 | 95,937 | 39 | 4.1 |
| Mural and mesangial | 16,939 | 4 | 2.4 | 0 | 9,366 | 5 | 5.3 | 1 | 12,519 | 3 | 2.4 |
| Fibroblast | 25,297 | 22 | 8.7 | 6 | 32,863 | 46 | 14.0 | 21 | 120,457 | 172 | 14.3 |
| Immune | 10,867 | 2 | 1.8 | 0 | 43,902 | 13 | 3.0 | 0 | 103,215 | 17 | 1.6 |
| Other | 4,781 | 0 | 0.0 | 0 | 358 | 0 | 0.0 | 0 | – | – | – |

*Supplementary Table S1, continued: B. EPO-positive cells in the dataset of PDGFRβ-positive cells, by cell type*

| Cell type | Cells | *EPO* ≥1 | ≥1 per 10,000 | Participants with *EPO* ≥1 |
|---|---|---|---|---|
| Immune | 2,083 | 4 | 19.2 | 2 |
| Fibroblasts | 6,964 | 10 | 14.4 | 5 |
| Pericytes | 3,789 | 4 | 10.6 | 3 |
| Myofibroblasts | 13,130 | 12 | 9.1 | 6 |
| Endothelial | 2,341 | 0 | 0.0 | 0 |
| Epithelial | 461 | 0 | 0.0 | 0 |
| Mesangial cells | 1,003 | 0 | 0.0 | 0 |
| Vascular smooth muscle cells | 499 | 0 | 0.0 | 0 |

*C. Fibroblast versus other nuclei in the single-nucleus datasets*

| Dataset | *EPO* counts | Fibroblast nuclei, *n*/*N* | Per 10,000 | Other nuclei, *n*/*N* | Per 10,000 | Rate ratio (95% CI) |
|---|---|---|---|---|---|---|
| Single-nucleus data from the KORNERSTONE cohort | One or more | 22/25,297 | 8.70 | 220/814,468 | 2.70 | 3.2 (2.0 to 5.0) |
| Single-nucleus data from the KORNERSTONE cohort | Two or more | 6/25,297 | 2.37 | 2/814,468 | 0.02 | 96.6 (17.3 to 978.5) |
| KPMP single-nucleus dataset | One or more | 46/32,863 | 14.00 | 201/582,306 | 3.45 | 4.1 (2.9 to 5.6) |
| KPMP single-nucleus dataset | Two or more | 21/32,863 | 6.39 | 8/582,306 | 0.14 | 46.5 (19.8 to 121.4) |

*D. Spatial dataset by cell type*

| Cell type | Cells | ≥1 *EPO* | ≥1 per 10,000 | Control probe mean per 10,000 | ≥2 *EPO* | ≥2 per 10,000 | Control probe largest ≥2 |
|---|---|---|---|---|---|---|---|
| Proximal tubule | 1,065,116 | 2,155 | 20.2 | 0.7 | 31 | 0.3 | 7 |
| Fibroblast | 826,184 | 2,251 | 27.2 | 0.3 | 684 | 8.3 | 2 |
| Endothelium | 598,379 | 1,451 | 24.2 | 0.4 | 86 | 1.4 | 3 |
| Thick ascending limb | 587,165 | 1,416 | 24.1 | 0.5 | 130 | 2.2 | 3 |
| Immune cell | 413,258 | 1,062 | 25.7 | 0.5 | 45 | 1.1 | 1 |
| Vascular smooth muscle | 269,809 | 515 | 19.1 | 0.4 | 30 | 1.1 | 2 |
| Injured proximal tubule | 241,315 | 574 | 23.8 | 0.7 | 3 | 0.1 | 1 |
| Principal cell | 230,758 | 665 | 28.8 | 0.7 | 7 | 0.3 | 1 |
| Intercalated cell A | 154,241 | 409 | 26.5 | 0.6 | 2 | 0.1 | 1 |
| Distal convoluted tubule | 149,462 | 326 | 21.8 | 0.5 | 4 | 0.3 | 1 |
| Connecting tubule | 147,442 | 346 | 23.5 | 0.6 | 3 | 0.2 | 1 |
| Adaptive TAL | 134,216 | 297 | 22.1 | 0.6 | 1 | 0.1 | 1 |
| Descending thin limb | 95,245 | 228 | 23.9 | 0.6 | 1 | 0.1 | 1 |
| Mesangial cell | 61,857 | 120 | 19.4 | 0.5 | 0 | 0.0 | 0 |
| Podocyte | 56,109 | 106 | 18.9 | 0.4 | 0 | 0.0 | 0 |
| Parietal epithelial cell | 37,213 | 88 | 23.6 | 0.7 | 1 | 0.3 | 1 |
| Intercalated cell B | 30,355 | 95 | 31.3 | 0.7 | 1 | 0.3 | 1 |
| Proliferating cell | 29,043 | 83 | 28.6 | 0.5 | 2 | 0.7 | 0 |

| Cell type | Cells | ≥1 *EPO* | ≥1 per 10,000 | Control probe mean per 10,000 | ≥2 *EPO* | ≥2 per 10,000 | Control probe largest ≥2 |
|---|---|---|---|---|---|---|---|
| Juxtaglomerular cell | 14,697 | 26 | 17.7 | 0.4 | 0 | 0.0 | 0 |
| Schwann cell | 7,867 | 19 | 24.2 | 0.7 | 0 | 0.0 | 0 |
| Adipocyte | 921 | 1 | 10.9 | 1.1 | 0 | 0.0 | 0 |

*E. KPMP single-nucleus dataset: fibroblast and VSM/P subclasses*

| Class | Subclass or region | Nuclei | *EPO* ≥1 | Per 10,000 (95% CI) | *EPO* ≥2 |
|---|---|---|---|---|---|
| Fibroblast | FIB | 14,189 | 29 | 20.4 (13.7 to 29.3) | 15 |
| Fibroblast | infFIB | 2,460 | 5 | 20.3 (6.6 to 47.4) | 3 |
| Fibroblast | M-FIB | 5,110 | 6 | 11.7 (4.3 to 25.5) | 2 |
| Fibroblast | dM-FIB | 3,148 | 3 | 9.5 (2.0 to 27.8) | 0 |
| Fibroblast | pvFIB | 2,499 | 2 | 8.0 (1.0 to 28.9) | 1 |
| VSM/P | VSMC | 2,829 | 2 | 7.1 (0.9 to 25.5) | 0 |
| VSM/P | MC | 1,472 | 1 | 6.8 (0.2 to 37.8) | 0 |
| VSM/P | VSMC/P | 3,863 | 2 | 5.2 (0.6 to 18.7) | 1 |
| Fibroblast | MYOF | 4,071 | 1 | 2.5 (0.1 to 13.7) | 0 |
| Fibroblast | dFIB | 1,386 | 0 | 0.0 (0.0 to 26.6) | 0 |
| VSM/P | dVSMC | 442 | 0 | 0.0 (0.0 to 83.1) | 0 |
| VSM/P | REN | 760 | 0 | 0.0 (0.0 to 48.4) | 0 |
| Fibroblast | All | 32,863 | 46 | 14.0 (10.2 to 18.7) | 21 |
| VSM/P | All | 9,366 | 5 | 5.3 (1.7 to 12.5) | 1 |
| Fibroblast | Region: cortex | 18,927 | 26 | 13.7 (9.0 to 20.1) | – |
| Fibroblast | Region: corticomedullary region | 5,866 | 16 | 27.3 (15.6 to 44.3) | – |
| Fibroblast | Region: medulla | 3,776 | 2 | 5.3 (0.6 to 19.1) | – |
| Fibroblast | Region: papilla | 4,123 | 2 | 4.9 (0.6 to 17.5) | – |

*F. Clustering*

| EPO-expressing cells | With another within 30 µm | Observed | Expected (95% range) | Observed/ expected | *P* |
|---|---|---|---|---|---|
| 667 | 431 | 64.6% | 3.7% (1.9 to 5.7) | 17.5 | <0.001 |

*Supplementary Table S1, continued: G. EPO transcripts in other cell types*

| Cell type | Cells | In contact, *n* (%) | Matched cells in contact, *n*/*N* | Matched in contact |
|---|---|---|---|---|
| Thick ascending limb | 130 | 62 (48%) | 9/1,300 | 0.7% |
| Endothelium | 86 | 54 (63%) | 3/860 | 0.3% |
| Immune cell | 45 | 20 (44%) | 0/450 | 0.0% |
| Proximal tubule | 31 | 10 (32%) | 1/310 | 0.3% |
| Vascular smooth muscle | 30 | 14 (47%) | 0/300 | 0.0% |
| Principal cell | 7 | 1 (14%) | 0/70 | 0.0% |
| Distal convoluted tubule | 4 | 0 (0%) | 0/40 | 0.0% |
| Connecting tubule | 3 | 0 (0%) | 0/30 | 0.0% |
| Injured proximal tubule | 3 | 1 (33%) | 0/30 | 0.0% |
| Proliferating cell | 2 | 0 (0%) | 0/20 | 0.0% |
| Intercalated cell A | 2 | 0 (0%) | 0/20 | 0.0% |
| Adaptive TAL | 1 | 1 (100%) | 0/10 | 0.0% |
| Parietal epithelial cell | 1 | 0 (0%) | 0/10 | 0.0% |
| Descending thin limb | 1 | 0 (0%) | 0/10 | 0.0% |
| Intercalated cell B | 1 | 0 (0%) | 0/10 | 0.0% |
| All | 347 | 163 (47%) | 13/3,470 | 0.4% |

*H. EPO transcripts in other cell types: location in the nucleus*

| Group | Cells | *EPO* transcripts | In the nucleus, *n* (%) |
|---|---|---|---|
| EPO-expressing fibroblasts | 684 | 3,745 | 1,556 (41.5%) |
| Other cells | 347 | 1,002 | 310 (30.9%) |

A, the participants of each dataset: composition by the dataset's own classification; age, eGFR and female sex as recorded for the participants with a value (KPMP datasets: age in 10-year bands, the band holding the median; eGFR recorded for 138 of the single-nucleus and 202 of the spatial participants, sex for 153 of the single-nucleus participants); the dataset of PDGFR*β*-positive cells holds no clinical values in its deposited data. Cells or nuclei, those analyzed (spots for the KPMP spatial transcriptomics dataset); fibroblasts, the fibroblasts of the spatial dataset and of the external Xenium dataset and the fibroblasts, myofibroblasts and pericytes of the dataset of PDGFR*β*-positive cells; EPO-positive, fibroblasts with two or more *EPO* transcripts (spatial dataset, external Xenium dataset), fibroblast nuclei or cells with an *EPO* count (single-cell datasets) and spots with an *EPO* count (KPMP spatial transcriptomics dataset), with the participants in parentheses (the samples holding such a spot for the KPMP spatial transcriptomics dataset). The 125 samples of the single-nucleus data from the KORNERSTONE cohort come from 125 participants, 8 of whom are also in the spatial dataset. Plasma, samples within 30 days of the biopsy; days between biopsy and blood median 0 (IQR 0, 0), largest 13.

B, cells or nuclei of each cell class with one or more and two or more *EPO* counts, per 10,000; the dataset of PDGFR*β*-positive cells by its own cell types.

C, the two single-nucleus datasets: nuclei with one or more and with two or more *EPO* counts among fibroblast nuclei and among all other nuclei (every other class), per 10,000, and the rate ratio of fibroblast versus other nuclei with its exact 95% interval conditional on the total of such nuclei.

D, the spatial dataset (154 participants) by cell type: cells with at least one and at least two *EPO* transcripts (quality value ≥20), the same counts for the 40 negative control probes (mean over probes for one or more, the largest probe for two or more). Fibroblasts with two or more *EPO* transcripts among those with one or more: 684 of 2,251 (30%), compared with 11 of 1,100 (1%) for the 40 negative control probes combined; Fisher's exact test, $P<0.001$.

E, nuclei of the KPMP single-nucleus dataset with one or more and two or more *EPO* counts in the subclasses of the fibroblast and the vascular smooth muscle and pericyte (VSM/P) classes of the KPMP kidney atlas, and the fibroblast nuclei by region of the sample (cortex, corticomedullary region, medulla and papilla; 32,692 of the 32,863 fibroblast nuclei, the others without a recorded region); per 10,000 nuclei with one or more counts, with exact (Clopper–Pearson) 95% intervals.

F, EPO-expressing cells in squares with a tissue zone with another EPO-expressing cell within 30 µm, against 1,000 random reassignments of EPO expression to the same number of fibroblasts of the same section and zone (expected, mean of the reassignments; 95% range, their 2.5th and 97.5th percentiles; *P*, one plus the reassignments at or above the observed value over 1,001).

G, the cells other than fibroblasts with two or more *EPO* transcripts by cell type: in contact, the cell boundary within 5 µm of the boundary of an EPO-expressing fibroblast; matched cells, ten cells of the same section and cell type without an *EPO* transcript and with a similar number of transcripts for each cell (45 of the cells lay in sections without an EPO-expressing fibroblast); odds ratio of contact 236 (126 to 441), logistic regression with errors clustered by participant.

H, *EPO* transcripts overlapping the nucleus in the other cells against EPO-expressing fibroblasts: odds ratio 0.63 (0.49 to 0.81), binomial model with errors clustered by participant.

–, not recorded or not in the dataset. AKI, acute kidney injury; C3GN, C3 glomerulonephritis; CI, confidence interval; CKD, chronic kidney disease; dFIB, degenerative fibroblast; DKD, diabetic kidney disease; dM-FIB, degenerative medullary fibroblast; DM-R, diabetes mellitus–resilient (KPMP enrollment category); dVSMC, degenerative vascular smooth muscle cell; eGFR, estimated glomerular filtration rate; EPO, erythropoietin; FIB, fibroblast; FSGS, focal segmental glomerulosclerosis; IgAN, IgA nephropathy; infFIB, inflammatory fibroblast; KPMP, Kidney Precision Medicine Project; M-FIB, medullary fibroblast; MC, mesangial cell; MCD, minimal change disease; MN, membranous nephropathy; MYOF, myofibroblast; pvFIB, perivascular fibroblast; REN, renin cell; TAL, thick ascending limb; VSMC, vascular smooth muscle cell; VSMC/P, vascular smooth muscle cell or pericyte.

**Supplementary Table S2.** EPO-expressing cells compared with matched fibroblasts

*A. Genes detected more or less often*

| Detected more often (25 of 101 genes) | | | | | | | Detected less often (25 of 124 genes) | | | | | | |
|---|---|---|---|---|---|---|---|---|---|---|---|---|---|
| Gene | Detected, % (expr./matched) | Log RR | *z* | *q* | Same zone log RR | Same zone *q* | Gene | Detected, % (expr./matched) | Log RR | *z* | *q* | Same zone log RR | Same zone *q* |
| *PTGIS* | 31.9/2.3 | 2.94 | 21.4 | <0.001 | 3.08 | <0.001 | *BGN* | 13.7/44.0 | −1.50 | −10.4 | <0.001 | −1.41 | <0.001 |
| *DPT* | 31.1/4.4 | 2.23 | 19.9 | <0.001 | 2.22 | <0.001 | *GPX3* | 10.4/24.0 | −0.99 | −10.0 | <0.001 | −0.83 | <0.001 |
| *H19* | 67.8/11.2 | 2.62 | 17.6 | <0.001 | 2.77 | <0.001 | *TINAGL1* | 4.2/17.3 | −1.57 | −10.0 | <0.001 | −1.46 | <0.001 |
| *OLFM1* | 29.8/3.5 | 2.42 | 16.8 | <0.001 | 2.49 | <0.001 | *AEBP1* | 8.2/23.2 | −1.19 | −9.1 | <0.001 | −1.05 | <0.001 |
| *NBL1* | 28.7/10.8 | 1.14 | 16.6 | <0.001 | 1.06 | <0.001 | *TNC* | 1.8/22.1 | −2.77 | −8.5 | <0.001 | −3.01 | <0.001 |
| *F8* | 14.6/0.3 | 4.02 | 15.6 | <0.001 | 4.15 | <0.001 | *WNT5A* | 0.6/5.4 | −2.29 | −8.1 | <0.001 | −2.28 | <0.001 |
| *VEGFA* | 24.6/2.5 | 2.48 | 15.3 | <0.001 | 2.58 | <0.001 | *FLNA* | 15.6/27.2 | −0.68 | −7.3 | <0.001 | −0.69 | <0.001 |
| *EPAS1* | 78.4/40.2 | 1.37 | 14.5 | <0.001 | 1.40 | <0.001 | *CXCL12* | 16.5/34.0 | −0.89 | −7.2 | <0.001 | −0.91 | <0.001 |
| *GREM2* | 13.2/0.7 | 3.07 | 11.8 | <0.001 | 2.98 | <0.001 | *VCAN* | 1.2/11.8 | −2.54 | −7.1 | <0.001 | −2.28 | <0.001 |
| *INSR* | 19.4/4.6 | 1.57 | 10.8 | <0.001 | 1.67 | <0.001 | *MYLK* | 8.6/17.4 | −0.80 | −6.8 | <0.001 | −0.84 | <0.001 |
| *ITIH5* | 18.4/5.6 | 1.31 | 9.9 | <0.001 | 1.27 | <0.001 | *ACTA2* | 15.1/30.7 | −0.91 | −6.3 | <0.001 | −0.76 | <0.001 |
| *NHERF2* | 18.1/7.7 | 0.93 | 9.9 | <0.001 | 0.78 | <0.001 | *CCN1* | 9.8/12.7 | −0.48 | −6.2 | <0.001 | −0.36 | 0.046 |
| *ASPN* | 19.0/6.2 | 1.26 | 9.6 | <0.001 | 1.34 | <0.001 | *MYH9* | 22.8/32.1 | −0.46 | −6.0 | <0.001 | −0.50 | <0.001 |
| *CDKN1C* | 10.8/3.5 | 1.20 | 8.6 | <0.001 | 1.08 | <0.001 | *LOXL1* | 2.3/8.3 | −1.35 | −5.9 | <0.001 | −1.33 | <0.001 |
| *ADM* | 14.6/3.0 | 1.67 | 8.4 | <0.001 | 1.74 | <0.001 | *ITGA1* | 6.9/16.5 | −1.00 | −5.7 | <0.001 | −0.98 | <0.001 |
| *ANGPTL2* | 15.6/4.7 | 1.30 | 8.3 | <0.001 | 1.36 | <0.001 | *CCDC80* | 1.9/7.6 | −1.47 | −5.6 | <0.001 | −1.25 | <0.001 |
| *IGF2* | 18.6/1.4 | 2.74 | 8.2 | <0.001 | 2.70 | <0.001 | *HSD17B2* | 0.7/5.1 | −2.02 | −5.2 | <0.001 | −2.15 | <0.001 |
| *RAPGEF5* | 16.4/4.7 | 1.35 | 8.1 | <0.001 | 1.35 | <0.001 | *PTGIR* | 1.5/5.8 | −1.45 | −5.2 | <0.001 | −1.38 | <0.001 |
| *ZNF395* | 9.1/3.4 | 1.04 | 8.0 | <0.001 | 1.04 | <0.001 | *ADRA2A* | 1.5/8.2 | −1.80 | −5.2 | <0.001 | −1.95 | <0.001 |
| *CCBE1* | 14.9/6.7 | 0.87 | 7.9 | <0.001 | 0.76 | <0.001 | *CSRP1* | 6.7/14.1 | −0.83 | −5.1 | <0.001 | −0.79 | <0.001 |
| *TMEM45A* | 5.3/1.4 | 1.37 | 7.6 | <0.001 | 1.50 | <0.001 | *PLXNB2* | 12.7/21.7 | −0.64 | −5.1 | <0.001 | −0.65 | <0.001 |
| *AOC3* | 33.3/21.4 | 0.57 | 7.5 | <0.001 | 0.59 | <0.001 | *IL13RA1* | 5.4/10.8 | −0.75 | −5.1 | <0.001 | −0.71 | <0.001 |
| *TRIB2* | 14.3/6.1 | 0.94 | 7.3 | <0.001 | 0.91 | <0.001 | *CCN2* | 10.7/19.4 | −0.84 | −5.0 | <0.001 | −0.77 | <0.001 |
| *EGFL7* | 4.1/1.1 | 1.37 | 7.0 | <0.001 | 1.35 | <0.001 | *PARM1* | 2.8/12.1 | −1.57 | −5.0 | <0.001 | −1.68 | <0.001 |
| *P4HA1* | 12.6/5.1 | 0.95 | 7.0 | <0.001 | 0.94 | <0.001 | *KLF4* | 0.4/1.6 | −1.41 | −5.0 | <0.001 | −1.24 | <0.001 |

*B. Hallmark gene sets in tissue sections*

| Gene set | Genes | Median *z* | *q* |
|---|---|---|---|
| Epithelial–mesenchymal transition | 75 | −2.21 | <0.001 |
| IL6 JAK STAT3 signaling | 40 | −1.43 | 0.003 |
| Interferon γ response | 78 | −1.04 | 0.023 |
| Oxidative phosphorylation | 43 | 0.27 | 0.023 |
| Inflammatory response | 64 | −1.10 | 0.023 |
| DNA repair | 35 | 0.05 | 0.023 |
| Allograft rejection | 56 | −1.19 | 0.033 |
| TNF-*α* signaling via NF-*κ*B | 76 | −1.05 | 0.033 |
| Fatty acid metabolism | 32 | 0.60 | 0.038 |

Comparison of 684 EPO-expressing cells of 81 participants with 6,840 matched fibroblasts (ten per case, drawn from the 30 fibroblasts of the same participant with the nearest log library size).

A, the genes at $q < 0.05$ in the cell-level complementary log–log model with participant fixed effects and standard errors clustered by participant (Benjamini–Hochberg over the 2,743 genes detected in at least 1% of the cells): the 25 genes with the largest absolute $z$ in each direction of the 101 detected more often and the 124 detected less often, ordered by absolute $z$; all genes are in Supplementary Data 2; percent of EPO-expressing (expr.) and matched fibroblasts with the gene detected; log RR, log rate ratio of detection; $z$, log RR over its standard error clustered by participant.

B, Hallmark sets at $q < 0.05$ in the rank-sum test of the $z$ of the set's genes compared with all other tested genes (Benjamini–Hochberg within the collection); median $z$ of the set's genes (negative, detected less often in EPO-expressing cells). Same zone, the same model with the ten fibroblasts of each EPO-expressing cell drawn from the same tissue zone of the same participant (667 EPO-expressing cells; the 17 in squares without a zone left out; Benjamini–Hochberg over the 2,743 genes fitted); RR, rate ratio; –, not fitted (separation). *COL1A1*, not among the genes at $q < 0.05$, was detected in 37.9% of EPO-expressing and 32.5% of matched cells ($q$ 0.538). All 2,743 genes and all sets are in Supplementary Data 2. EPO, erythropoietin.

**Supplementary Table S3.** Genes of EPO-positive fibroblasts in single nuclei and profile-positive cells

*A. Genes of the 70 tested in the spatial dataset*

| | | Spatial dataset | | | |
|---|---|---|---|---|---|
| Gene | Nuclei, combined *z* | EPO-expressing, % | Matched fibroblasts, % | Log rate ratio | *q* |
| *PTGIS* | 8.8 | 31.9 | 2.3 | 2.94 | <0.001 |
| *DPT* | 5.4 | 31.1 | 4.4 | 2.23 | <0.001 |
| *H19* | 6.0 | 67.8 | 11.2 | 2.62 | <0.001 |
| *OLFM1* | 4.9 | 29.8 | 3.5 | 2.42 | <0.001 |
| *F8* | 10.6 | 14.6 | 0.3 | 4.02 | <0.001 |
| *VEGFA* | 9.5 | 24.6 | 2.5 | 2.48 | <0.001 |
| *GREM2* | 9.5 | 13.2 | 0.7 | 3.07 | <0.001 |
| *INSR* | 5.6 | 19.4 | 4.6 | 1.57 | <0.001 |
| *ITIH5* | 5.8 | 18.4 | 5.6 | 1.31 | <0.001 |
| *ANGPTL2* | 3.7 | 15.6 | 4.7 | 1.30 | <0.001 |
| *IGF2* | 4.9 | 18.6 | 1.4 | 2.74 | <0.001 |
| *EGFL7* | 6.2 | 4.1 | 1.1 | 1.37 | <0.001 |
| *P4HA1* | 4.9 | 12.6 | 5.1 | 0.95 | <0.001 |
| *NPR3* | 4.7 | 6.6 | 2.6 | 0.98 | <0.001 |
| *EDNRB* | 4.6 | 20.9 | 10.5 | 0.81 | <0.001 |
| *COLEC12* | 4.9 | 7.9 | 3.2 | 0.96 | <0.001 |
| *MEG3* | 5.5 | 21.6 | 10.5 | 0.81 | <0.001 |
| *ESR2* | 4.3 | 5.4 | 2.3 | 0.87 | 0.004 |
| *COL23A1* | 5.6 | 3.5 | 1.4 | 0.95 | 0.022 |
| *PDE4B* | 4.3 | 3.1 | 1.6 | 0.65 | 0.033 |
| *VIPR2* | 3.8 | 2.8 | 1.2 | 0.84 | 0.038 |
| *DNPEP* | 3.9 | 2.3 | 1.5 | 0.45 | 0.252 |
| *ABCC9* | 4.2 | 16.8 | 14.0 | 0.21 | 0.272 |
| *FHL1* | 3.9 | 2.2 | 3.1 | −0.38 | 0.399 |
| *F2R* | 4.5 | 9.8 | 7.5 | 0.27 | 0.417 |
| *CCDC137* | 4.0 | 0.7 | 1.1 | −0.38 | 0.727 |
| *SRCAP* | 3.8 | 3.7 | 3.9 | −0.07 | 0.897 |
| *CCL11* | 4.5 | 1.5 | 1.6 | −0.10 | 0.915 |
| *APLF* | 4.0 | 1.3 | 1.4 | −0.08 | 0.916 |

*Supplementary Table S3, continued: B. Genes of EPO-positive fibroblast nuclei, datasets combined*

| Gene | Log rate ratio | *z* | *q* | Datasets | On spatial panel |
|---|---|---|---|---|---|
| *F8* | 2.71 | 10.6 | <0.001 | KPMP, PDGFR*β*, KORNERSTONE | Yes |
| *VEGFA* | 1.93 | 9.5 | <0.001 | KPMP, PDGFR*β*, KORNERSTONE | Yes |
| *GREM2* | 2.77 | 9.5 | <0.001 | KPMP, PDGFR*β*, KORNERSTONE | Yes |
| *PTGIS* | 1.84 | 8.8 | <0.001 | KPMP, PDGFR*β*, KORNERSTONE | Yes |
| *PDE8B* | 1.91 | 8.7 | <0.001 | KPMP, PDGFR*β* | No |
| *HIF1A-AS3* | 2.17 | 7.4 | <0.001 | KPMP | No |
| *FRMPD4* | 4.11 | 7.0 | <0.001 | KPMP | No |
| *LOXL3* | 2.45 | 6.6 | <0.001 | KPMP, PDGFR*β* | No |
| *EGFL7* | 1.96 | 6.2 | <0.001 | KPMP, PDGFR*β*, KORNERSTONE | Yes |
| *NEBL* | 1.39 | 6.0 | <0.001 | KPMP, PDGFR*β*, KORNERSTONE | Yes |
| *H19* | 1.48 | 6.0 | <0.001 | KPMP, PDGFR*β*, KORNERSTONE | Yes |
| *TGFBI* | 1.20 | 6.0 | <0.001 | KPMP, PDGFR*β* | No |
| *ITIH5* | 1.01 | 5.8 | <0.001 | KPMP, PDGFR*β*, KORNERSTONE | Yes |
| *COL23A1* | 1.20 | 5.6 | <0.001 | KPMP, PDGFR*β*, KORNERSTONE | Yes |
| *INSR* | 0.89 | 5.6 | <0.001 | KPMP, PDGFR*β*, KORNERSTONE | Yes |
| *MEG3* | 0.90 | 5.5 | <0.001 | KPMP, PDGFR*β*, KORNERSTONE | Yes |
| *NPAS2* | 1.23 | 5.4 | <0.001 | KPMP, PDGFR*β* | No |
| *DPT* | 1.35 | 5.4 | <0.001 | KPMP, PDGFR*β*, KORNERSTONE | Yes |
| *COLEC12* | 0.94 | 4.9 | <0.001 | KPMP, PDGFR*β*, KORNERSTONE | Yes |
| *OLFM1* | 1.35 | 4.9 | <0.001 | KPMP, PDGFR*β*, KORNERSTONE | Yes |
| *IGF2* | 1.52 | 4.9 | <0.001 | PDGFR*β* | Yes |
| *P4HA1* | 0.93 | 4.9 | <0.001 | KPMP, PDGFR*β*, KORNERSTONE | Yes |
| *PITPNM2* | 1.43 | 4.7 | 0.002 | KPMP, PDGFR*β* | No |
| *NPR3* | 0.99 | 4.7 | 0.002 | KPMP, PDGFR*β*, KORNERSTONE | Yes |
| *SMIM3* | 0.93 | 4.7 | 0.002 | KPMP, PDGFR*β* | No |
| *PIK3R6* | 5.09 | 4.6 | 0.003 | KPMP | No |
| *EDNRB* | 0.87 | 4.6 | 0.003 | KPMP, PDGFR*β*, KORNERSTONE | Yes |
| *CCL11* | 1.35 | 4.5 | 0.003 | KPMP, PDGFR*β* | Yes |
| *GPSM1* | 1.58 | 4.5 | 0.003 | KPMP, PDGFR*β* | No |
| *F2R* | 0.94 | 4.5 | 0.004 | KPMP, PDGFR*β* | Yes |
| *AC078881.1* | 1.57 | 4.4 | 0.006 | KPMP | No |
| *PLOD2* | 0.95 | 4.4 | 0.006 | KPMP, PDGFR*β* | No |
| *ESR2* | 0.89 | 4.3 | 0.007 | KPMP, PDGFR*β*, KORNERSTONE | Yes |
| *RFLNB* | 1.90 | 4.3 | 0.008 | KPMP, PDGFR*β* | No |
| *ZNF131* | 0.84 | 4.3 | 0.009 | KPMP, PDGFR*β* | No |
| *AQP5* | 2.75 | 4.3 | 0.009 | KPMP, PDGFR*β* | Yes |
| *PDE4B* | 1.18 | 4.3 | 0.009 | KPMP, PDGFR*β* | Yes |
| *AC093772.1* | 1.19 | 4.2 | 0.010 | KPMP, PDGFR*β* | No |
| *ABCC9* | 0.66 | 4.2 | 0.012 | KPMP, PDGFR*β*, KORNERSTONE | Yes |

| Gene | Log rate ratio | $z$ | $q$ | Datasets | On spatial panel |
|---|---|---|---|---|---|
| *ZNF730* | 2.84 | 4.2 | 0.012 | KPMP | No |
| *ADH1B* | 0.78 | 4.2 | 0.012 | KPMP, PDGFR$\beta$ | No |
| *ATP11A* | 0.95 | 4.2 | 0.012 | KPMP, PDGFR$\beta$ | No |
| *POP7* | 1.28 | 4.1 | 0.014 | KPMP, PDGFR$\beta$ | No |
| *AC026904.2* | 2.59 | 4.1 | 0.015 | KPMP, PDGFR$\beta$ | No |
| *PI4KB* | 1.12 | 4.1 | 0.015 | KPMP, PDGFR$\beta$ | No |
| *RXFP1* | 0.98 | 4.1 | 0.015 | KPMP, PDGFR$\beta$, KORNERSTONE | Yes |
| *ALPK2* | 1.89 | 4.1 | 0.016 | KPMP | No |
| *CCDC137* | 1.45 | 4.0 | 0.018 | KPMP, PDGFR$\beta$, KORNERSTONE | Yes |
| *EFNB1* | 1.17 | 4.0 | 0.019 | KPMP, PDGFR$\beta$ | No |
| *APLF* | 1.02 | 4.0 | 0.023 | KPMP, KORNERSTONE | Yes |
| *HIST1H2BD* | 2.82 | 4.0 | 0.023 | KPMP | No |
| *DNPEP* | 0.94 | 3.9 | 0.024 | KPMP, PDGFR$\beta$, KORNERSTONE | Yes |
| *FAM43A* | 1.23 | 3.9 | 0.024 | KPMP, PDGFR$\beta$ | No |
| *FHL1* | 0.77 | 3.9 | 0.024 | KPMP, PDGFR$\beta$, KORNERSTONE | Yes |
| *TENM3* | 0.87 | 3.9 | 0.024 | KPMP, PDGFR$\beta$ | No |
| *WIPF3* | 2.39 | 3.9 | 0.024 | KPMP | No |
| *PDE4C* | 1.03 | 3.9 | 0.027 | KPMP, PDGFR$\beta$ | No |
| *MEG8* | 0.84 | 3.9 | 0.027 | KPMP, PDGFR$\beta$ | No |
| *SRCAP* | 1.06 | 3.8 | 0.035 | KPMP, PDGFR$\beta$, KORNERSTONE | Yes |
| *MIR155HG* | 2.98 | 3.8 | 0.037 | PDGFR$\beta$ | No |
| *BAMBI* | 1.09 | 3.8 | 0.039 | KPMP, PDGFR$\beta$ | No |
| *APCDD1L* | 2.62 | 3.8 | 0.039 | KPMP, PDGFR$\beta$ | No |
| *SH2D2A* | 2.69 | 3.8 | 0.039 | PDGFR$\beta$ | Yes |
| *VIPR2* | 1.16 | 3.8 | 0.040 | KPMP, PDGFR$\beta$, KORNERSTONE | Yes |
| *ELF4* | 2.68 | 3.8 | 0.041 | KPMP | Yes |
| *AC092902.2* | 2.04 | 3.7 | 0.042 | PDGFR$\beta$ | No |
| *SPIN4* | 2.30 | 3.7 | 0.042 | PDGFR$\beta$ | No |
| *ARHGAP21* | 0.69 | 3.7 | 0.043 | KPMP, PDGFR$\beta$ | No |
| *COL6A1* | 0.71 | 3.7 | 0.043 | KPMP, PDGFR$\beta$ | No |
| *ANGPTL2* | 1.04 | 3.7 | 0.045 | KPMP, PDGFR$\beta$, KORNERSTONE | Yes |
| *NAV2* | −0.92 | −4.1 | 0.012 | KPMP, PDGFR$\beta$, KORNERSTONE | Yes |
| *SYNPO2* | −0.96 | −4.2 | 0.011 | KPMP, PDGFR$\beta$, KORNERSTONE | Yes |

*Supplementary Table S3, continued: C. Gene-set tests*

| Group or direction | Collection | Gene set | Hits/set genes or set genes | Fold or median *z* | *q* |
|---|---|---|---|---|---|
| *Over-representation among the 70 genes detected more often* | | | | | |
| Imprinted genes | Geneimprint | Imprinted or predicted | 6/127 | 10.9 | <0.001 |
| | Geneimprint | Imprinted | 4/70 | 13.2 | <0.001 |
| Signaling by G protein-coupled receptors | Reactome | Signaling by GPCR | 10/348 | 6.7 | 0.003 |
| | GO BP | G protein-coupled receptor signaling pathway | 9/380 | 5.5 | 0.019 |
| | Reactome | Peptide ligand binding receptors | 4/57 | 16.3 | 0.035 |
| Cyclic nucleotide metabolic process | GO BP | Cyclic purine nucleotide metabolic process | 4/29 | 32.0 | 0.007 |
| | GO BP | Cyclic nucleotide metabolic process | 4/30 | 30.9 | 0.007 |
| | GO BP | Cyclic nucleotide catabolic process | 3/14 | 49.6 | 0.017 |
| | GO BP | cAMP metabolic process | 3/17 | 40.9 | 0.025 |
| | GO BP | Negative regulation of cAMP/PKA signal transduction | 3/26 | 26.7 | 0.048 |
| Collagen formation | GO BP | Extracellular matrix organization | 7/160 | 10.1 | 0.007 |
| | GO BP | External encapsulating structure organization | 7/160 | 10.1 | 0.007 |
| | GO BP | Extracellular structure organization | 7/161 | 10.1 | 0.007 |
| | Reactome | Collagen formation | 5/69 | 16.8 | 0.008 |
| | GO BP | Collagen fibril organization | 4/52 | 17.8 | 0.026 |
| | Reactome | Collagen biosynthesis and modifying enzymes | 4/54 | 17.2 | 0.035 |
| Regulation of body fluid levels | GO BP | Regulation of body fluid levels | 7/195 | 8.3 | 0.014 |
| Leukocyte chemotaxis | GO BP | Myeloid leukocyte migration | 4/52 | 17.8 | 0.026 |
| | GO BP | Leukocyte chemotaxis | 4/61 | 15.2 | 0.040 |
| Muscle cell development | GO BP | Striated muscle cell development | 4/62 | 14.9 | 0.040 |
| | GO BP | Cardiac muscle cell development | 3/27 | 25.7 | 0.049 |
| Broad (more than 1,000 genes) | GO BP | Anatomical structure morphogenesis | 16/1,065 | 3.5 | 0.007 |
| | GO BP | Signaling | 26/2,839 | 2.1 | 0.026 |
| | GO BP | Signal transduction | 25/2,673 | 2.2 | 0.026 |
| | GO BP | Cell communication | 26/2,862 | 2.1 | 0.027 |
| | GO BP | Multicellular organismal process | 27/3,122 | 2.0 | 0.040 |
| | GO BP | Regulation of multicellular organismal process | 19/1,833 | 2.4 | 0.049 |
| *Hallmark rank tests of the combined z of all tested genes* | | | | | |
| Lower | Hallmark | Oxidative phosphorylation | 198 | −0.06 | <0.001 |
| | Hallmark | MYC targets v1 | 189 | 0.03 | <0.001 |
| | Hallmark | Coagulation | 97 | −0.14 | 0.001 |

*Supplementary Table S3, continued: D. Transcription factor and pathway activity*

| | Nuclei, datasets combined | | | | Spatial dataset | | | | |
|---|---|---|---|---|---|---|---|---|---|
| Factor or pathway | Target or response genes | Activity (*t*) | *q* | Own combined *z* | Response genes on the panel | Activity (*t*) | *q* | Activity, same zone | *q*, same zone |
| *Transcription factors (CollecTRI), q < 0.05* | | | | | | | | | |
| PRRX1 | 5 | 5.21 | <0.001 | −1.0 | – | – | – | – | – |
| NFATC4 | 26 | 4.92 | <0.001 | 2.2 | – | – | – | – | – |
| HLF | 13 | 4.38 | 0.001 | 0.2 | – | – | – | – | – |
| HDGF | 6 | 3.49 | 0.027 | 0.0 | – | – | – | – | – |
| MYT1 | 5 | 3.48 | 0.027 | – | – | – | – | – | – |
| FOXM1 | 56 | 3.44 | 0.028 | – | – | – | – | – | – |
| PEG3 | 7 | 3.42 | 0.029 | 1.7 | – | – | – | – | – |
| HOXB7 | 15 | 3.37 | 0.030 | 0.3 | – | – | – | – | – |
| ARNT | 48 | 3.31 | 0.036 | 0.3 | – | – | – | – | – |
| HDAC3 | 16 | 3.29 | 0.036 | 0.8 | – | – | – | – | – |
| EPAS1 | 71 | 3.26 | 0.038 | 3.3 | – | – | – | – | – |
| FOXP3 | 32 | 3.20 | 0.044 | – | – | – | – | – | – |
| HIF1A | 344 | 3.16 | 0.046 | 1.1 | – | – | – | – | – |
| JUNB | 60 | 3.15 | 0.046 | −0.1 | – | – | – | – | – |
| TEAD4 | 6 | 3.11 | 0.050 | −0.1 | – | – | – | – | – |
| HIPK2 | 8 | −3.18 | 0.046 | 2.3 | – | – | – | – | – |
| RXRB | 10 | −3.39 | 0.030 | 0.6 | – | – | – | – | – |
| FOXO4 | 22 | −3.68 | 0.015 | – | – | – | – | – | – |
| HOXC6 | 10 | −3.69 | 0.015 | −0.2 | – | – | – | – | – |
| MTA3 | 10 | −3.86 | 0.009 | 0.7 | – | – | – | – | – |
| RUNX1 | 132 | −4.17 | 0.003 | 0.5 | – | – | – | – | – |
| ZMYND8 | 6 | −4.51 | <0.001 | −0.9 | – | – | – | – | – |
| SMAD5 | 27 | −4.80 | <0.001 | 0.0 | – | – | – | – | – |
| HIF3A | 5 | −6.36 | <0.001 | −1.5 | – | – | – | – | – |
| *Signaling pathways (PROGENy), all 14* | | | | | | | | | |
| Hypoxia | 465 | 6.28 | <0.001 | – | 128 | 6.33 | <0.001 | 7.57 | <0.001 |
| MAPK | 450 | 2.93 | 0.016 | – | 117 | −1.12 | 0.406 | −0.97 | 0.493 |
| PI3K | 394 | 1.83 | 0.120 | – | 74 | 0.98 | 0.456 | 0.90 | 0.493 |
| TRAIL | 228 | 1.82 | 0.120 | – | 49 | 0.65 | 0.553 | 0.61 | 0.581 |
| EGFR | 422 | 1.68 | 0.144 | – | 89 | −1.27 | 0.398 | −0.95 | 0.493 |
| Estrogen | 422 | 0.69 | 0.625 | – | 125 | −0.74 | 0.553 | −0.97 | 0.493 |
| VEGF | 370 | 0.57 | 0.664 | – | 80 | −1.24 | 0.398 | −0.86 | 0.493 |
| p53 | 350 | 0.34 | 0.793 | – | 86 | −0.71 | 0.553 | −0.20 | 0.842 |
| WNT | 332 | 0.01 | 0.992 | – | 77 | −0.52 | 0.603 | −0.68 | 0.581 |
| Androgen | 425 | −1.59 | 0.158 | – | 125 | −1.21 | 0.398 | −1.60 | 0.339 |
| NF-$\kappa$B | 392 | −1.86 | 0.120 | – | 128 | −1.98 | 0.207 | −1.55 | 0.339 |

| Factor or pathway | Nuclei, datasets combined: Target or response genes | Activity ($t$) | $q$ | Own combined $z$ | Spatial dataset: Response genes on the panel | Activity ($t$) | $q$ | Activity, same zone | $q$, same zone |
|---|---|---|---|---|---|---|---|---|---|
| JAK–STAT | 400 | −1.91 | 0.120 | – | 115 | −1.89 | 0.207 | −1.58 | 0.339 |
| TNF-$\alpha$ | 392 | −2.23 | 0.089 | – | 124 | −1.77 | 0.216 | −1.43 | 0.359 |
| TGF-$\beta$ | 404 | −3.21 | 0.009 | – | 130 | −3.99 | <0.001 | −3.24 | 0.008 |

*D, continued. Response genes of the hypoxia and TGF-β pathways in the nuclei*

| Pathway | Ranks | Response genes and their contribution (weight × combined $z$) |
|---|---|---|
| Hypoxia | 1–20 | *VEGFA* 101.30; *NDRG1* 69.40; *PLOD2* 59.42; *P4HA1* 53.88; *TMEM45A* 47.52; *ADM* 43.37; *PDK1* 39.78; *IGFBP3* 32.62; *GBE1* 26.10; *PGM1* 24.93; *MXI1* 22.26; *VLDLR* 19.39; *LDHA* 17.48; *LOX* 16.98; *BNIP3* 15.82; *EGLN3* 15.55; *ZNF395* 15.33; *ARRDC3* 14.92; *PGK1* 14.35; *ANKRD37* 14.22 |
| TGF-$\beta$ | 1–20 | *ID3* −31.82; *PMEPA1* −17.54; *MFAP4* −14.45; *TAGLN* −12.65; *CCDC80* −12.49; *ID1* −11.25; *PDE8B* −11.10; *LRRC32* −10.06; *CAVIN2* −10.03; *TGFBR2* −9.50; *RXFP1* −9.28; *VCAN* −9.04; *UACA* −8.66; *SULF1* −8.66; *FRMPD4* −8.48; *DACT1* −8.31; *CEBPD* −8.04; *MAMLD1* −7.77; *UNC5B* −7.76; *LMCD1* −7.29 |

*E. Published hypoxia and EPO-producing cell lists*

| List | Genes of the 70 | Genes |
|---|---|---|
| Hallmark hypoxia | 3 | *VEGFA*, *TGFBI*, *P4HA1* |
| Reactome cellular response to hypoxia | 1 | *VEGFA* |
| Reactome regulation of gene expression by HIF | 1 | *VEGFA* |
| GO response to hypoxia (with descendants) | 3 | *VEGFA*, *PTGIS*, *PLOD2* |
| Norn cell markers of the mouse (Kragesteen et al.) | 3 | *ITIH5*, *F2R*, *COL6A1* |
| In none of these lists | 62 | – |

*F. Genes tied to EPO-producing cells in earlier work*

| Gene | Reference | Datasets | Log rate ratio | $z$ | $P$ | $q$ |
|---|---|---|---|---|---|---|
| *EPAS1* | [S17, S20] | KPMP, PDGFR$\beta$, KORNERSTONE | 0.57 | 3.3 | <0.001 | 0.119 |
| *PDGFRB* | [S19, S20] | KPMP, PDGFR$\beta$, KORNERSTONE | −0.46 | −2.5 | 0.014 | 0.394 |
| *PDGFRA* | [S16] | KPMP, PDGFR$\beta$, KORNERSTONE | −0.20 | −1.2 | 0.218 | 0.873 |
| *CXCL12* | [S16] | KPMP, PDGFR$\beta$, KORNERSTONE | −0.36 | −1.8 | 0.071 | 0.660 |
| *ACTA2* | [S18, S19] | KPMP, PDGFR$\beta$, KORNERSTONE | −0.31 | −1.6 | 0.109 | 0.748 |
| *EGLN1* | [S19, S20] | KPMP, PDGFR$\beta$, KORNERSTONE | 0.12 | 0.5 | 0.641 | 0.964 |
| *EGLN3* | [S19, S20] | KPMP, PDGFR$\beta$, KORNERSTONE | 0.60 | 1.3 | 0.182 | 0.841 |
| *TCF21* | [S16, S20] | KPMP, PDGFR$\beta$, KORNERSTONE | 0.08 | 0.3 | 0.741 | 0.976 |
| *CEBPD* | [S20] | KPMP | 0.65 | 2.4 | 0.016 | 0.420 |
| *ADM* | [S20] | KPMP, PDGFR$\beta$ | 0.84 | 2.4 | 0.015 | 0.405 |
| *RGS4* | [S20] | PDGFR$\beta$ | 0.54 | 0.7 | 0.474 | 0.948 |

*G. Profile-positive cells*

| Dataset or analysis | Measure | Value | Population |
|---|---|---|---|
| Spatial dataset | Profile-positive cells | 8,991 | 1.1% of 826,184 fibroblasts; 153 participants |
| Spatial dataset, by zone | Cortical labyrinth | 4.1 per 1,000 | 1,715 of 418,751 fibroblasts; 154 biopsies |
| Spatial dataset, by zone | Zone of straight tubules | 11.6 per 1,000 | 3,631 of 313,019 fibroblasts; 154 biopsies |
| Spatial dataset, by zone | Deeper medulla | 65.6 per 1,000 | 3,001 of 45,716 fibroblasts; 97 biopsies |
| KPMP fibroblast nuclei | Profile-positive on the 20 genes of the other two datasets, no *EPO* count | 2,078 | 6.3% of the fibroblast nuclei |
| KPMP fibroblast nuclei | EPO-positive nuclei, median percentile of the score | 94 | – |

A, the 29 of the 70 genes detected more often in EPO-positive fibroblast nuclei (B) that were tested in the spatial dataset (Supplementary Figure S2b; 34 of the 70 are on the panel, and genes detected in fewer than 1% of the fibroblasts were not tested): the 21 detected more often in EPO-expressing cells at $q < 0.05$ first; combined $z$ of the nuclei; percent of EPO-expressing and matched fibroblasts with the gene detected, log rate ratio and $q$ of the clustered model of Supplementary Table S2.

B, genes at combined $q < 0.05$ in EPO-positive versus matched fibroblast nuclei of the same participant in the KPMP single-nucleus dataset (45 nuclei of 29 participants, whole transcriptome), the single-nucleus data from the KORNERSTONE cohort (KORNERSTONE; 22 nuclei, 5,070 genes) and the dataset of PDGFR$\beta$-positive cells (PDGFR$\beta$; 26 mesenchymal cells of 6 patients), combined with inverse-variance weights over the 16,217 genes tested in any dataset.

C, every Gene Ontology biological process, Reactome and imprinted-gene set at $q < 0.05$ in the hypergeometric test of the 70 genes detected more often, with all tested genes as background ($q$ within each collection; hits over set genes among the tested genes), grouped as in the Results, sets with more than 1,000 tested genes listed as broad (no Hallmark or KEGG set reached $q < 0.05$); and the Hallmark sets at $q < 0.05$ in the Mann–Whitney test of the combined $z$ of their members compared with all other tested genes.

D, univariate linear model of the combined $z$ of all tested genes on the signed CollecTRI weights of a factor's targets, or on the weights of the 500 response genes of a PROGENy pathway (activity, the $t$ of the slope; $q$ over the 644 factors with at least five targets and over the 14 pathways); own combined $z$, the factor's gene in the combined comparison; spatial dataset, the same pathway model on the $z$ of the clustered model of Supplementary Table S2 (response genes on the panel), and on the $z$ of the comparison with fibroblasts of the same tissue zone (same zone).

D, continued, the response genes of the hypoxia and TGF-$\beta$ pathways in the nuclei with their contribution to the activity, the PROGENy weight times the combined $z$ of the gene: the 20 response genes with the largest contribution in the direction of each activity (the hypoxia activity was positive and the TGF-$\beta$ activity negative; the first five of each are named in Figure 2d); all response genes are in Supplementary Data 2.

E, the 70 genes in published hypoxia gene sets and in the Norn cell markers of the mouse mapped to human genes.

F, genes tied to EPO-producing cells in the references, their rows of B for all 16,217 genes ($q$ over all genes).

G, fibroblasts of the spatial dataset without an *EPO* transcript and with a profile score at or above the median of the EPO-expressing cells (profile-positive cells); profile-positive cells per 1,000 fibroblasts of each tissue zone (zone of the 50-µm square holding the cell; zones as in Supplementary Table S6 B, no test); KPMP fibroblast nuclei scored on the genes chosen in the other two single-cell datasets, profile-positive nuclei with no *EPO* count and a score at or above the median of the EPO-positive nuclei, and the median percentile of the EPO-positive nuclei among the fibroblast nuclei without an *EPO* count. All results are in Supplementary Data 2.

EPO, erythropoietin; HIF, hypoxia-inducible factor; KPMP, Kidney Precision Medicine Project.

**Supplementary Table S4.** Location of EPO-expressing cells

*A. Neighbors within 30 μm (%)*

| Neighbors | EPO-expressing | Other fibroblasts | Median difference | Higher | *P* | *q* |
|---|---|---|---|---|---|---|
| Proximal tubule | 19.0 | 15.7 | 2.4 | 42/81 | 0.173 | 0.190 |
| Injured proximal tubule | 1.7 | 4.3 | −3.0 | 14/81 | <0.001 | <0.001 |
| Thick ascending limb | 33.1 | 9.7 | 25.6 | 64/81 | <0.001 | <0.001 |
| Endothelium | 11.8 | 11.6 | −0.4 | 39/81 | 0.587 | 0.587 |
| Vascular smooth muscle | 2.5 | 6.1 | −4.6 | 13/81 | <0.001 | <0.001 |
| Immune cells | 3.7 | 10.9 | −5.7 | 9/81 | <0.001 | <0.001 |
| Fibroblasts | 18.8 | 24.7 | −6.8 | 20/81 | <0.001 | <0.001 |
| Distal convoluted tubule | 2.6 | 1.8 | −1.1 | 9/81 | <0.001 | <0.001 |
| Connecting tubule | 0.3 | 2.0 | −1.6 | 3/81 | <0.001 | <0.001 |
| Principal cells | 1.7 | 2.9 | −2.2 | 12/81 | <0.001 | <0.001 |
| Cells within 30 μm, *n* | 16.4 | 15.9 | 1.1 | 53/81 | 0.013 | 0.015 |
| *Immune cell types adjacent to EPO-expressing and matched fibroblasts* | | | | | | |
| resident macrophages | 1.0 | 2.2 | −1.5 | 11/81 | <0.001 | <0.001 |
| immune cells of unresolved type | 0.4 | 2.0 | −1.3 | 6/81 | <0.001 | <0.001 |
| CD4 and regulatory T cells | 0.6 | 1.7 | −1.0 | 5/81 | <0.001 | <0.001 |
| inflammatory myeloid cells | 0.5 | 1.6 | −1.0 | 8/81 | <0.001 | <0.001 |
| cytotoxic T cells | 0.3 | 0.9 | −0.5 | 7/81 | <0.001 | <0.001 |
| plasma cells | 0.5 | 0.9 | −0.3 | 12/81 | 0.002 | 0.003 |
| dendritic cells | 0.2 | 0.9 | −0.6 | 3/81 | <0.001 | <0.001 |
| B cells | 0.0 | 0.7 | −0.2 | 0/81 | <0.001 | <0.001 |
| NK-like cells | 0.1 | 0.4 | −0.2 | 6/81 | <0.001 | <0.001 |
| mast cells | 0.1 | 0.2 | 0.0 | 6/81 | <0.001 | <0.001 |
| neutrophils | 0.0 | 0.1 | 0.0 | 3/81 | <0.001 | <0.001 |

*B. Distance to the nearest cell (μm)*

| Nearest cell | EPO-expressing | Other fibroblasts | Median difference | Higher | *P* | *q* |
|---|---|---|---|---|---|---|
| Glomerulus | 1,155.8 | 1,162.5 | 23.3 | 42/81 | 0.957 | 0.957 |
| Proximal tubule | 33.7 | 87.9 | −17.4 | 13/81 | <0.001 | <0.001 |
| Injured proximal tubule | 74.9 | 69.9 | −1.5 | 38/81 | 0.673 | 0.770 |
| Thick ascending limb | 27.0 | 72.4 | −52.0 | 7/81 | <0.001 | <0.001 |
| Extraglomerular endothelium | 18.1 | 22.3 | −7.0 | 25/81 | <0.001 | <0.001 |
| Vascular smooth muscle | 51.0 | 48.1 | 0.1 | 41/81 | 0.587 | 0.770 |
| Juxtaglomerular cell | 485.1 | 433.2 | 27.6 | 43/81 | 0.456 | 0.729 |
| Immune cell | 44.9 | 34.1 | 9.5 | 51/81 | <0.001 | <0.001 |
| Thick ascending limb, median of participant medians | 12.0 | 47.2 | −24.8 (−38.4 to −19.8) | 9/81 | <0.001 | – |

*Supplementary Table S4, continued: C. KPMP spatial transcriptomics dataset (Visium)*

| Signature | EPO-positive spot (SD) | Same direction | Adjacent spots (SD) | Same direction |
|---|---|---|---|---|
| Thick ascending limb | 0.36 (0.27 to 0.48) | 104/137 | 0.28 (0.20 to 0.37) | 104/137 |
| Proximal tubule | 0.16 (0.09 to 0.24) | 78/137 | 0.10 (0.04 to 0.17) | 85/137 |
| Injured proximal tubule | −0.02 (−0.08 to 0.02) | 81/137 | −0.02 (−0.07 to 0.01) | 78/137 |
| Immune cells | −0.06 (−0.13 to −0.01) | 83/137 | −0.07 (−0.12 to −0.03) | 80/137 |

*D. External Xenium dataset*

| Neighbors | Fibroblasts | Samples | Cells | Mean of sample means, % | Median of sample means, % |
|---|---|---|---|---|---|
| Thick ascending limb | EPO-expressing cells | 11 | 172 | 21.1 | 7.6 |
| Thick ascending limb | Other fibroblasts | 11 | 95,202 | 4.9 | 3.1 |
| Thick ascending limb, with the injured state | EPO-expressing cells | 11 | 172 | 22.7 | 8.6 |
| Thick ascending limb, with the injured state | Other fibroblasts | 11 | 95,202 | 8.8 | 7.5 |

A and B, means over the 81 participants with an EPO-expressing cell of the participant means of EPO-expressing and of other fibroblasts; neighbor proportions are percentages of the 20 nearest cells within 30 µm; distances to the nearest cell of each kind in µm; Wilcoxon signed-rank test, Benjamini–Hochberg *q* within the proportions and within the distances; for the distal convoluted tubule the mean proportion is higher near EPO-expressing cells because a few participants have a high proportion, while the median difference is negative and most participants have a lower proportion. The immune cell types of A compare the neighbors of EPO-expressing cells with those of their matched fibroblasts (column Other fibroblasts), with *q* over all neighbor measures of that comparison (Supplementary Table S5 and Supplementary Data 3). The last row of B gives the medians over participants of the participant medians (the values of Results), the median difference with its 95% confidence interval and the sign test.

C, KPMP spatial transcriptomics dataset (Visium): difference of the standardized signature score of spots with an *EPO* UMI and of their six adjacent spots from depth-matched spots of the same sample; samples with the same direction over evaluable samples.

D, external Xenium dataset: proportion of thick ascending limb cells (without and with the cells of the injured state) among the 30-µm neighbors of fibroblasts with two or more *EPO* counts and of the other fibroblasts; mean and median over samples of the sample means; no test. EPO, erythropoietin; KPMP, Kidney Precision Medicine Project.

**Supplementary Table S5.** Tubular neighbors of EPO-expressing cells

*A. Neighbors*

| Neighbors | EPO-expressing | Matched fibroblasts | Mean difference | Participants higher | *q* |
|---|---|---|---|---|---|
| Tubular neighbors in a healthy state | 91.3 | 66.9 | 24.4 | 75/79 | <0.001 |
| Adaptive state (of thick ascending limb) | 7.9 | 38.3 | −30.3 | 3/71 | <0.001 |
| S3 segment (of proximal tubule) | 36.1 | 21.8 | 14.3 | 47/69 | <0.001 |
| Injured state (of proximal tubule) | 11.5 | 31.3 | −19.8 | 8/69 | <0.001 |

*B. Adjacent thick ascending limb*

| Measure | Partici-pants | TAL cells beside EPO-expressing | TAL cells beside matched | EPO-expressing | Matched fibroblasts | *q* |
|---|---|---|---|---|---|---|
| *UMOD* detected, % | 69 | 2,814 | 21,315 | 99.4 | 98.2 | 0.108 |
| *KCNJ1* detected, % | 69 | 2,814 | 21,315 | 33.5 | 31.4 | 0.799 |
| *ATP1A1* detected, % | 69 | 2,814 | 21,315 | 86.1 | 85.1 | 0.987 |
| Oxidative phosphorylation score | 69 | 2,814 | 21,315 | 0.054 | 0.053 | 0.799 |
| Hypoxia pathway activity | 69 | 2,814 | 21,315 | 6.55 | 6.66 | 0.799 |

A, participant means over the 30-µm neighbors (the 20 nearest cells within 30 µm) of the EPO-expressing cells and of their matched fibroblasts, proportions in percent (tubular neighbors in a healthy state; the adaptive state among thick ascending limb neighbors; the S3 segment and the injured state among proximal tubule neighbors, as defined for the proximal tubule of the spatial dataset); participants higher, participants whose EPO-expressing cells had the larger value (Wilcoxon signed-rank test, Benjamini–Hochberg *q* over all neighbor measures of Supplementary Data 3). All results are in Supplementary Data 3. B, thick ascending limb (TAL) cells in a healthy state within 30 µm of the EPO-expressing cells and within 30 µm of their fibroblasts matched in the same participant and tissue zone (receivers with at least one such cell: 581 EPO-expressing and 3,801 matched cells), TAL cells near both left out; participant means compared by the Wilcoxon signed-rank test over the participants with both groups, Benjamini–Hochberg *q* over the five measures; *SLC12A1* and *ATP1B1* are not on the panel. Oxidative phosphorylation score, mean normalized expression of the 46 panel genes of the Hallmark set; hypoxia pathway activity, PROGENy response genes on the panel (151). EPO, erythropoietin.

**Supplementary Table S6.** EPO-expressing cells and the surrounding tissue within biopsies

*A. Features of the surrounding tissue*

| Feature of the surrounding 150-µm square | All squares: Rate ratio per SD (95% CI) | All squares: *P* | Zone of straight tubules: Rate ratio per SD (95% CI) | Zone of straight tubules: *P* |
|---|---|---|---|---|
| Proportion of immune cells | 0.20 (0.12 to 0.31) | <0.001 | 0.21 (0.12 to 0.37) | <0.001 |
| Inflammatory signaling of fibroblasts | 0.54 (0.43 to 0.69) | <0.001 | 0.55 (0.42 to 0.74) | <0.001 |
| TNF-*α* pathway | 0.60 (0.45 to 0.80) | <0.001 | 0.59 (0.42 to 0.82) | 0.002 |
| NF-*κ*B pathway | 0.62 (0.48 to 0.81) | <0.001 | 0.62 (0.46 to 0.84) | 0.002 |
| JAK–STAT pathway | 0.60 (0.48 to 0.74) | <0.001 | 0.63 (0.48 to 0.83) | <0.001 |
| Contractile program | 0.37 (0.29 to 0.47) | <0.001 | 0.48 (0.35 to 0.65) | <0.001 |
| Proportion of injured proximal tubule cells | 0.55 (0.40 to 0.74) | <0.001 | 0.55 (0.39 to 0.77) | <0.001 |
| Collagen matrix program | 1.06 (0.59 to 1.90) | 0.844 | 1.08 (0.53 to 2.19) | 0.834 |
| *Proportion of immune cells, contractile program, proportion of injured PT cells and collagen matrix program in one model, all squares* | | | | |
| Proportion of immune cells | 0.29 (0.18 to 0.46) | <0.001 | – | – |
| Contractile program | 0.37 (0.27 to 0.50) | <0.001 | – | – |
| Proportion of injured proximal tubule cells | 0.60 (0.47 to 0.77) | <0.001 | – | – |
| Collagen matrix program | 1.73 (1.11 to 2.68) | 0.015 | – | – |

*B. Tissue zones*

| Zone | Biopsies | Squares | TAL cells, % of cells | Fibroblasts | EPO-expressing cells | Per 1,000 fibroblasts | Rate ratio within biopsies (95% CI) |
|---|---|---|---|---|---|---|---|
| Cortical labyrinth | 154 | 146,045 | 2.8 | 418,751 | 52 | 0.12 | 1 (reference) |
| Zone of straight tubules | 154 | 101,331 | 27.1 | 313,019 | 615 | 1.96 | 11.6 (7.1 to 19.1) |
| Deeper medulla | 97 | 14,359 | 24.3 | 45,716 | 0 | 0.00 | – |

*C. Deciles of the features of A*

| Decile | Proportion of immune cells: Fibro-blasts | Proportion of immune cells: EPO-expr. | Inflammatory signaling: Fibro-blasts | Inflammatory signaling: EPO-expr. | Contractile program: Fibro-blasts | Contractile program: EPO-expr. | Proportion of injured PT cells: Fibro-blasts | Proportion of injured PT cells: EPO-expr. | Collagen matrix program: Fibro-blasts | Collagen matrix program: EPO-expr. |
|---|---|---|---|---|---|---|---|---|---|---|
| 1 | 22,867 | 83 | 25,550 | 89 | 25,574 | 111 | 26,069 | 46 | 23,158 | 64 |
| 2 | 25,677 | 98 | 31,048 | 130 | 33,297 | 81 | 28,490 | 101 | 29,276 | 62 |
| 3 | 26,719 | 120 | 33,685 | 81 | 35,094 | 91 | 26,961 | 78 | 32,105 | 73 |
| 4 | 28,173 | 69 | 36,649 | 80 | 36,143 | 95 | 27,206 | 77 | 33,800 | 87 |
| 5 | 29,637 | 69 | 37,581 | 60 | 38,123 | 56 | 28,935 | 56 | 34,703 | 71 |
| 6 | 31,609 | 39 | 38,876 | 57 | 38,014 | 71 | 31,050 | 96 | 36,451 | 57 |
| 7 | 34,789 | 79 | 38,335 | 57 | 38,610 | 55 | 34,649 | 64 | 37,395 | 51 |
| 8 | 40,019 | 60 | 38,383 | 28 | 38,836 | 53 | 39,387 | 70 | 39,301 | 76 |
| 9 | 48,146 | 29 | 37,711 | 28 | 37,953 | 24 | 47,995 | 45 | 41,219 | 53 |
| 10 | 66,302 | 0 | 36,120 | 36 | 32,294 | 9 | 63,196 | 13 | 46,530 | 52 |

*D. Tissue zones without the thick ascending limb in the straight segments*

| Zone | Biopsies | Squares | Fibroblasts | EPO-expressing cells | Per 1,000 fibroblasts | Rate ratio within biopsies (95% CI) |
|---|---|---|---|---|---|---|
| Cortical labyrinth | 154 | 183,080 | 535,826 | 300 | 0.56 | 1 (reference) |
| Zone of straight tubules | 154 | 64,384 | 197,730 | 362 | 1.83 | 2.12 (1.11 to 4.03) |
| Deeper medulla | 94 | 14,105 | 44,742 | 0 | 0.00 | – |

*E. Thick ascending limb cells among the neighbors within zones*

| Zone | Participants | EPO-expressing cells | EPO-expressing, % | Other fibroblasts, % | Higher/lower | *P* |
|---|---|---|---|---|---|---|
| *Zones of B* | | | | | | |
| Cortical labyrinth | 26 | 52 | 7.1 | 2.7 | 7/19 | 0.727 |
| Zone of straight tubules | 71 | 615 | 43.4 | 24.2 | 62/9 | <0.001 |
| *Zones of D* | | | | | | |
| Cortical labyrinth | 53 | 300 | 29.7 | 8.2 | 38/15 | <0.001 |
| Zone of straight tubules | 56 | 362 | 43.1 | 23.1 | 51/5 | <0.001 |

*F. Profile-positive cells by decile of the proportion of immune cells*

| Decile | Immune cells, median % | Fibroblasts | Profile-positive | Per 1,000 (95% CI) | EPO-expressing | Per 1,000 (95% CI) |
|---|---|---|---|---|---|---|
| 1 | 0.0 | 22,867 | 161 | 7.04 (6.04 to 8.21) | 83 | 3.63 (2.93 to 4.50) |
| 2 | 1.0 | 25,677 | 178 | 6.93 (5.99 to 8.02) | 98 | 3.82 (3.13 to 4.65) |
| 3 | 1.7 | 26,719 | 174 | 6.51 (5.62 to 7.55) | 120 | 4.49 (3.76 to 5.37) |
| 4 | 2.4 | 28,173 | 161 | 5.71 (4.90 to 6.66) | 69 | 2.45 (1.94 to 3.10) |
| 5 | 3.2 | 29,637 | 193 | 6.51 (5.66 to 7.49) | 69 | 2.33 (1.84 to 2.95) |
| 6 | 4.3 | 31,609 | 181 | 5.73 (4.95 to 6.62) | 39 | 1.23 (0.90 to 1.69) |
| 7 | 5.8 | 34,789 | 199 | 5.72 (4.98 to 6.57) | 79 | 2.27 (1.82 to 2.83) |
| 8 | 8.0 | 40,019 | 207 | 5.17 (4.52 to 5.92) | 60 | 1.50 (1.17 to 1.93) |
| 9 | 12.2 | 48,146 | 170 | 3.53 (3.04 to 4.10) | 29 | 0.60 (0.42 to 0.86) |
| 10 | 23.8 | 66,302 | 146 | 2.20 (1.87 to 2.59) | 0 | 0.00 (0.00 to 0.06) |

A, Poisson models of the EPO-expressing cells of 118,196 50-µm squares (participants with an EPO-expressing cell in such squares, 77; 646 EPO-expressing cells) with the fibroblasts of the square as exposure, participant fixed effects, log(1 + proximal tubule cells of the 150-µm block) and standard errors clustered by participant; each feature alone, per SD of the squares, over all squares and over the 46,925 squares of the zone of straight tubules (69 participants, 587 EPO-expressing cells), and the proportion of immune cells, the contractile program, the proportion of injured proximal tubule cells and the collagen matrix program in one model (inflammatory signaling not in this model). Inflammatory signaling of fibroblasts, mean over the fibroblasts other than EPO-expressing cells of the block of the mean standardized PROGENy scores of the TNF-$\alpha$, NF-$\kappa$B and JAK–STAT pathways (Supplementary Methods); the three pathways also alone; –, not fitted.

B, every 50-µm square holding a fibroblast placed in a zone by the tubular segments of its 150-µm block (Supplementary Methods; 15,089 squares with 17 EPO-expressing cells had no zone); rate ratio versus the cortical labyrinth from one Poisson model with the zone, participant fixed effects, the fibroblasts as exposure and errors clustered by participant (150,716 squares of the 80 participants with an EPO-expressing cell); TAL cells, the proportion of thick ascending limb cells among all cells of the zone's squares, pooled over sections; –, no EPO-expressing cell in the zone, no estimate (also in D).

C, the squares of A (all squares) by decile of each feature: fibroblasts and EPO-expressing (expr.) cells per decile, the rates of Figure 3e. CI, confidence interval; EPO, erythropoietin; PT, proximal tubule; TAL, thick ascending limb.

D, the zones of B with the thick ascending limb left out of the straight segments (proximal tubule S3, thin limb and collecting duct only; Supplementary Methods); rate ratio as in B (149,187 squares of the 79 participants with an EPO-expressing cell).

E, proportion of thick ascending limb cells among the 30-µm neighbors of EPO-expressing cells and of fibroblasts without an *EPO* transcript of the same zone, participant means (participants with an EPO-expressing cell in the zone), number of participants with a higher/lower proportion around EPO-expressing cells, Wilcoxon signed-rank *P*; the deeper medulla held EPO-expressing cells in 2 participants.

F, the squares and deciles of the proportion of immune cells of C: fibroblasts without an *EPO* transcript whose profile score reached the median of the EPO-expressing cells (profile-positive) and EPO-expressing cells per 1,000 fibroblasts, Wilson 95% CI; profile-positive cells per SD of the proportion of immune cells, model of A over the squares of the 150 participants with a profile-positive cell, rate ratio 0.73 (0.68 to 0.78) (EPO-expressing cells, 0.20 (0.12 to 0.31)).

**Supplementary Table S7.** EPO-expressing cells by eGFR, tissue measures, diagnosis and hemoglobin

*A. All patients, with diagnosis*

| Exposure | *n* | Rate ratio (95% CI) |
|---|---|---|
| eGFR, per 10 mL/min/1.73 $m^2$ | 130 | 1.22 (1.06 to 1.41) |
| With the proportion of injured proximal tubule cells: eGFR | 130 | 1.02 (0.87 to 1.20) |
| With the proportion of injured proximal tubule cells: the proportion, per SD | 130 | 0.40 (0.25 to 0.64) |

*B. By diagnosis*

| Exposure | *n* | Rate ratio (95% CI) |
|---|---|---|
| IgA nephropathy: eGFR, per 10 mL/min/1.73 $m^2$ | 60 | 1.23 (1.01 to 1.51) |
| DKD: eGFR, per 10 mL/min/1.73 $m^2$ | 28 | 0.93 (0.66 to 1.33) |

*C. Diagnoses*

| Exposure | *n* | Rate ratio (95% CI) |
|---|---|---|
| IgA nephropathy versus living donors | 154 | 0.77 (0.30 to 1.99) |
| DKD versus living donors | 154 | 0.73 (0.22 to 2.46) |
| MCD versus living donors | 154 | 0.24 (0.08 to 0.73) |
| MN versus living donors | 154 | 0.58 (0.17 to 2.02) |
| FSGS versus living donors | 154 | 0.68 (0.11 to 4.25) |
| DKD versus IgA nephropathy | 88 | 0.87 (0.37 to 2.07) |

*D. Hemoglobin*

| Exposure | *n* | Rate ratio (95% CI) |
|---|---|---|
| All patients: hemoglobin, per g/dL | 130 | 0.81 (0.65 to 1.02) |
| eGFR ≥60: hemoglobin, per g/dL | 104 | 0.75 (0.58 to 0.95) |
| eGFR <60: hemoglobin, per g/dL | 26 | 1.10 (0.64 to 1.92) |

Negative binomial models of the EPO-expressing cells of a participant with the number of fibroblasts as offset and age, sex, eGFR and the proportion of thick ascending limb cells among the section's cells as covariates; models over more than one diagnosis also hold the diagnosis. A, the proportion of injured proximal tubule cells (per SD) added to the eGFR model. B, the eGFR model within each of the two largest diagnoses. C, each diagnosis versus living donors in one model over all participants, and DKD versus IgA nephropathy. D, patients by kidney function (hemoglobin for all 130). EPO-expressing cells, fibroblasts with two or more *EPO* transcripts. CI, confidence interval; DKD, diabetic kidney disease; eGFR, estimated glomerular filtration rate; EPO, erythropoietin; FSGS, focal segmental glomerulosclerosis; MCD, minimal change disease; MN, membranous nephropathy.

**Supplementary Table S8.** Plasma EPO and EPO-expressing cells

| Exposure | *n* | Estimate (95% CI) |
|---|---|---|
| EPO-expressing cell density (SD per SD) | 100 | 0.30 (0.13 to 0.46) |

Plasma EPO (SD) within 30 days of the biopsy in patients with a spatial section, per SD of the density of EPO-expressing cells (cells per $mm^2$ of tissue, log); least squares with HC3 errors, age, sex, eGFR, plate and diagnosis. EPO-expressing cells, fibroblasts with two or more *EPO* transcripts. CI, confidence interval; eGFR, estimated glomerular filtration rate; EPO, erythropoietin.

### Supplementary Figures

#### Supplementary Figure S1

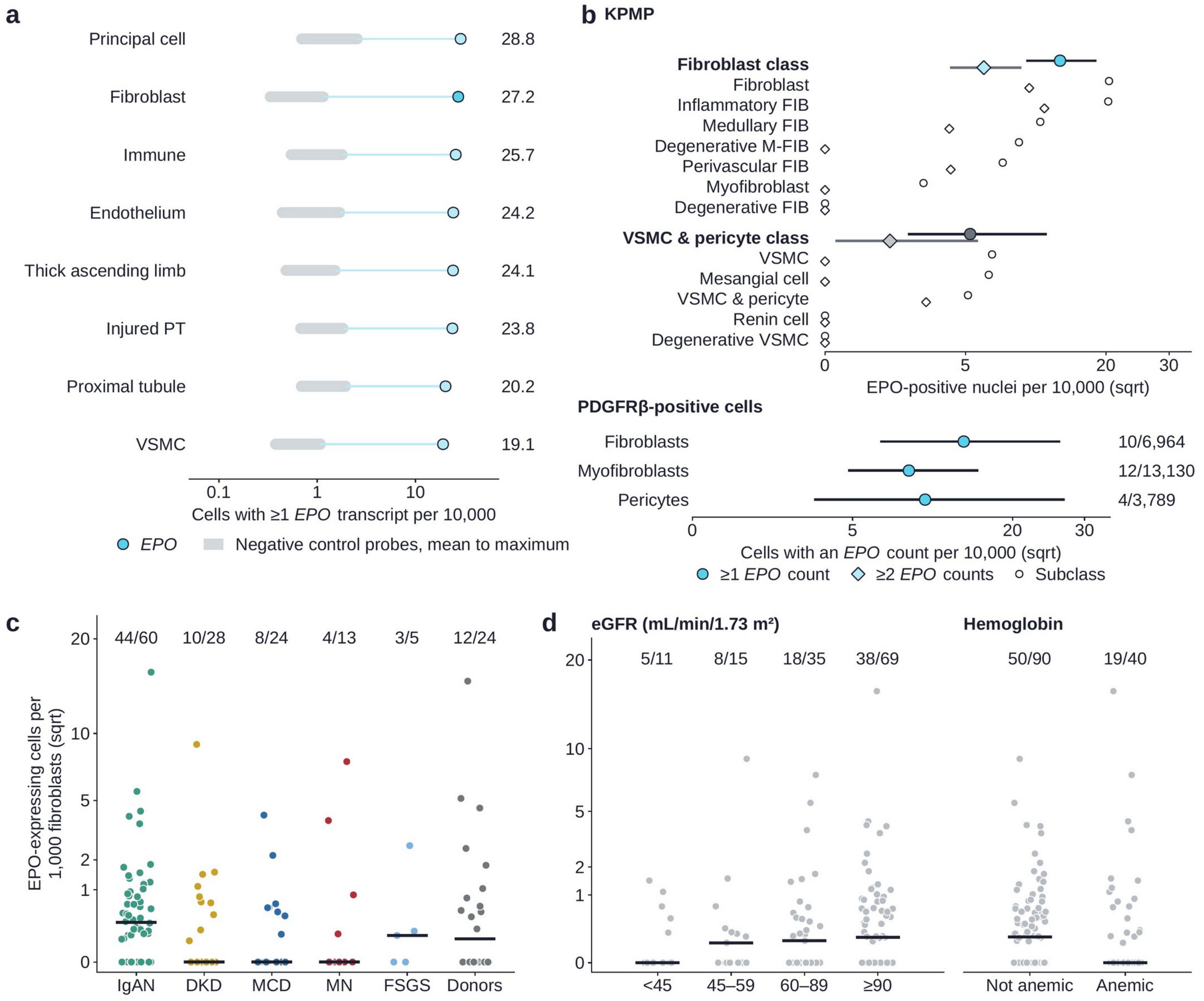

**Supplementary Figure S1. EPO-positive cells by cell type and by patient group.** **a**, Spatial dataset: cells with one or more *EPO* transcripts per 10,000 cells by cell type (logarithmic axis); gray band, the 40 negative control probes of the panel (mean to maximum). **b**, KPMP single-nucleus dataset: nuclei with one or more (circles) and two or more (diamonds) *EPO* counts per 10,000 nuclei in the fibroblast and the vascular smooth muscle and pericyte classes (filled, cyan for the fibroblast class and gray for the vascular smooth muscle and pericyte class, with exact 95% confidence intervals) and in their subclasses (open); and the dataset of PDGFR$\beta$-positive cells: cells with an *EPO* count per 10,000 fibroblasts, myofibroblasts and pericytes (exact 95% confidence intervals); square-root axes. **c**, Spatial dataset: EPO-expressing cells per 1,000 fibroblasts by diagnosis (points, participants; bars, medians; numbers, participants with an EPO-expressing cell over all participants). **d**, The same for the patients by eGFR group and by anemia. DKD, diabetic kidney disease; FIB, fibroblast; FSGS, focal segmental glomerulosclerosis; IgAN, IgA nephropathy; KPMP, Kidney Precision Medicine Project; M-FIB, medullary fibroblast; MCD, minimal change disease; MN, membranous nephropathy; PT, proximal tubule; VSMC, vascular smooth muscle cell.

**Supplementary Figure S2**

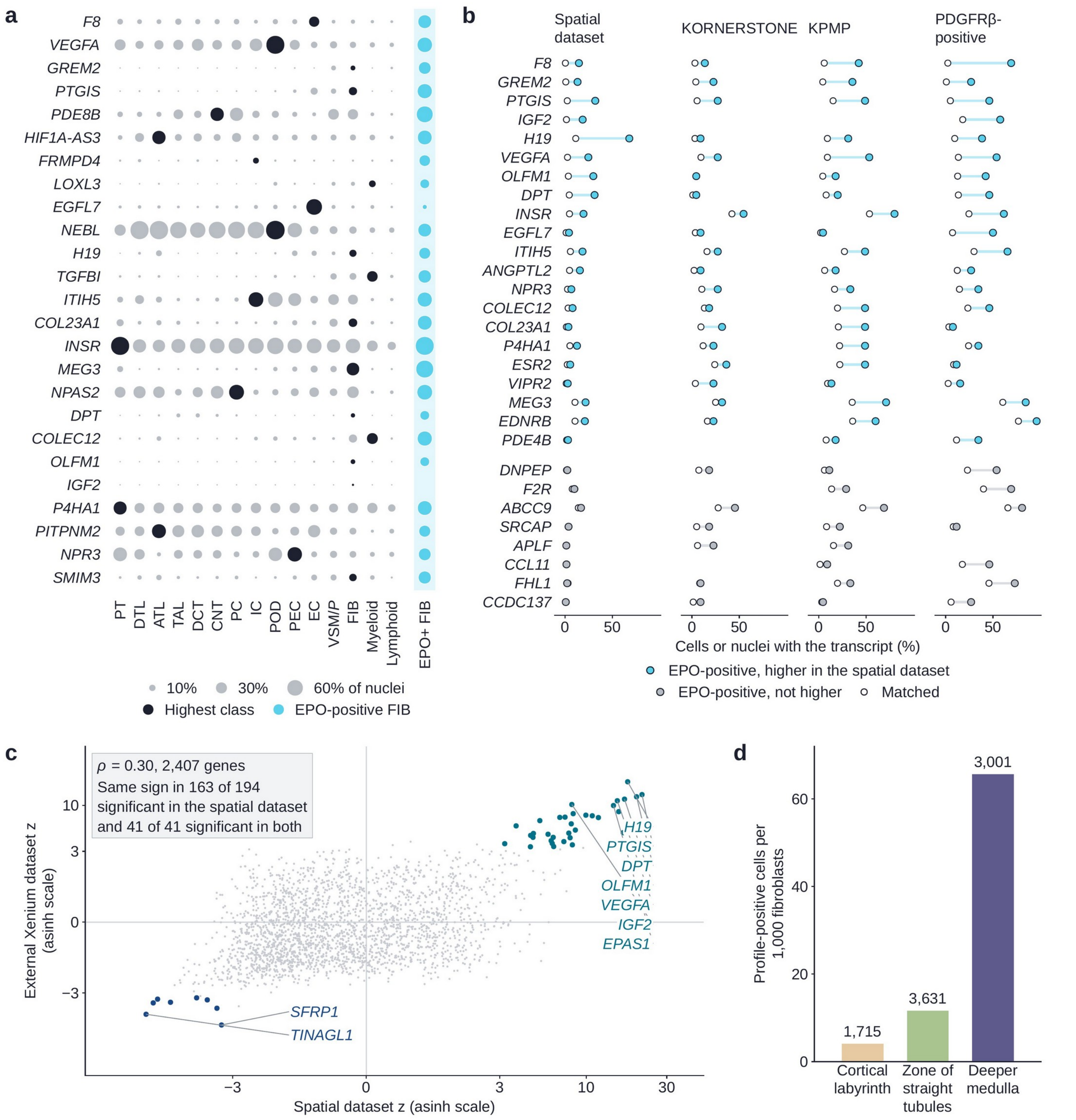

**Supplementary Figure S2. Genes of EPO-positive cells across datasets.** **a**, KPMP single-nucleus dataset: proportion of nuclei in which each of the 25 genes of Table 2 was detected, by cell class (dot area) and in the 46 EPO-positive fibroblast nuclei (cyan column); black, the class with the highest proportion for each gene; classes with fewer than 1,000 nuclei not shown. **b**, The 29 of the 70 genes detected more often in EPO-positive nuclei that were tested in the spatial dataset: percent of EPO-positive (filled) and of matched (open) cells or nuclei in which the gene was detected, in the spatial dataset and in the three single-cell datasets; top, the 21 genes detected more often in the spatial dataset ($q < 0.05$; cyan); bottom, the eight that did not differ (gray). **c**, *z* of EPO-expressing versus matched fibroblasts in the spatial dataset and in the external Xenium dataset for the 2,407 shared genes (inverse hyperbolic sine axes); colored, $q < 0.05$ in both datasets (teal, higher; blue, lower in EPO-expressing cells); labels, the six genes detected more often and the two detected less often in both datasets with the largest $|z|$ in the external Xenium dataset, and *EPAS1* and *VEGFA*, which the text names; gray box, Spearman $\rho$ and agreement of sign. **d**, Spatial dataset: profile-positive cells (fibroblasts without an *EPO* transcript that expressed the gene profile of EPO-positive fibroblasts) per 1,000 fibroblasts by tissue zone (numbers, cells). ATL, ascending thin limb; CNT, connecting tubule; DCT, distal convoluted tubule; DTL, descending thin limb; EC, endothelial cell; FIB, fibroblast; IC, intercalated cell; KPMP, Kidney Precision Medicine Project; PC, principal cell; PEC, parietal epithelial cell; POD, podocyte; PT, proximal tubule; TAL, thick ascending limb; VSM/P, vascular smooth muscle cell or pericyte.

Supplementary Figure S3

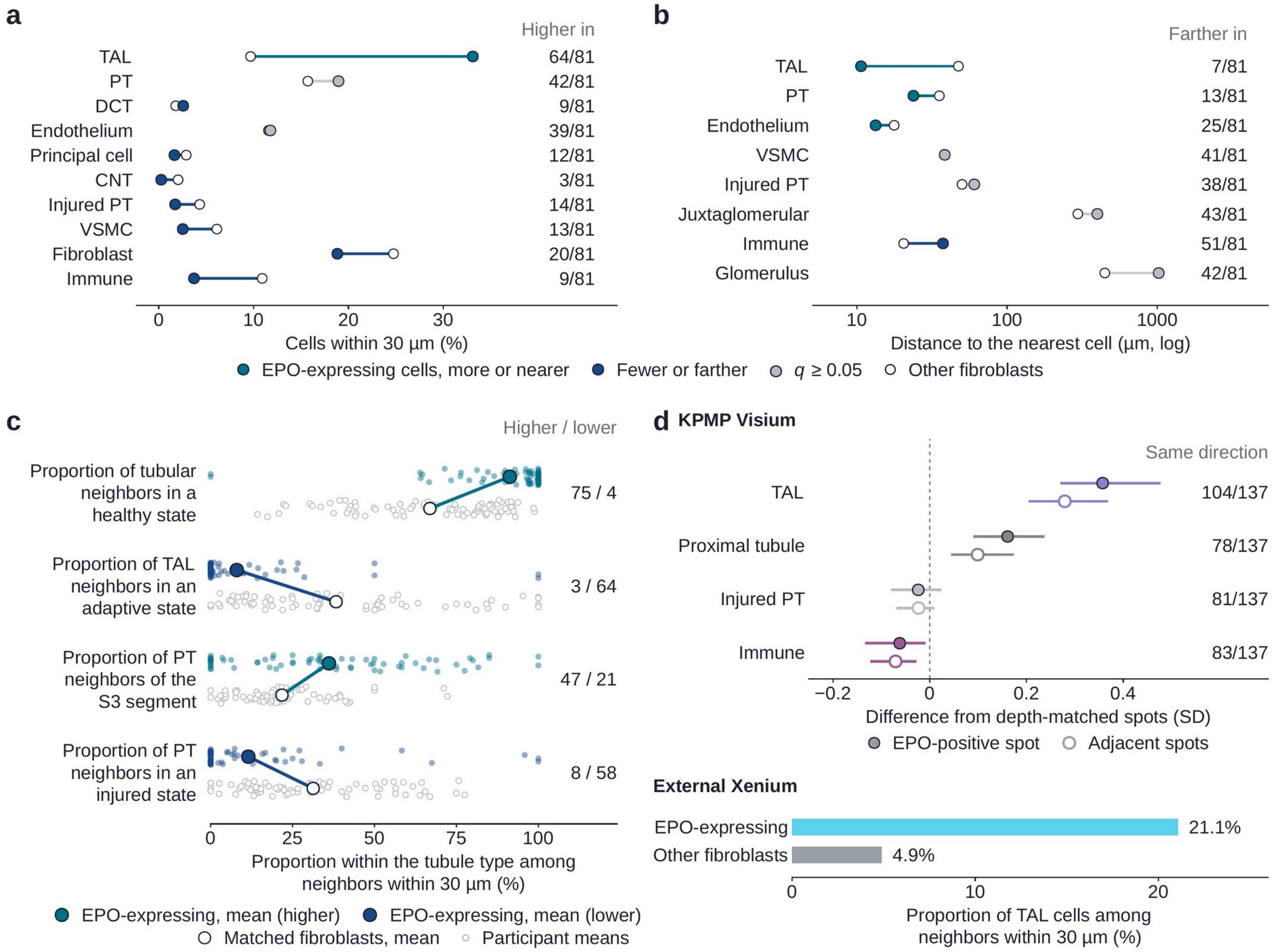

**Supplementary Figure S3. Neighborhood of EPO-expressing cells. a**, Spatial dataset: proportion of each cell class among the 20 nearest cells within 30 µm of EPO-expressing cells (filled) and of other fibroblasts (open), means across participants; dark cyan, more frequent near EPO-expressing cells; blue, less frequent; gray, $q \geq 0.05$; color gives the direction in most participants (for the distal convoluted tubule the mean is higher but the proportion was lower in most participants); numbers, participants with a higher proportion near EPO-expressing cells. **b**, Distance to the nearest cell of each kind (logarithmic axis), with the same marks; numbers, participants in whom the cell was located farther from EPO-expressing cells. **c**, Proportion of tubular neighbors in a healthy state, of thick ascending limb neighbors in an adaptive state, of proximal tubule neighbors of the S3 segment and of proximal tubule neighbors in an injured state, within 30 µm of EPO-expressing cells (filled) and of matched fibroblasts (open), means across participants; numbers, participants with a higher or a lower value around EPO-expressing cells; small points, participant means; dark cyan, higher and blue, lower around EPO-expressing cells. **d**, KPMP spatial transcriptomics dataset (Visium): difference of the thick ascending limb, proximal tubule, injured proximal tubule and immune signature scores of spots with an *EPO* count (filled) and of their six adjacent spots (open) from depth-matched spots of the same sample (SD; 95% confidence intervals from resamples of participants; colored, interval excluding 0; numbers, samples with the same direction over evaluable samples); and external Xenium dataset: proportion of thick ascending limb cells among the 30-µm neighbors of fibroblasts with two or more *EPO* counts and of other fibroblasts (means of the 11 samples with such cells). CNT, connecting tubule; DCT, distal convoluted tubule; KPMP, Kidney Precision Medicine Project; PT, proximal tubule; TAL, thick ascending limb; VSMC, vascular smooth muscle cell.
